# Contracting COVID-19: A Longitudinal Investigation of the Impact of Beliefs and Knowledge

**DOI:** 10.1101/2021.04.15.21255556

**Authors:** Courtney A. Moore, Benjamin C. Ruisch, Javier A. Granados Samayoa, Shelby T. Boggs, Jesse T. Ladanyi, Russell H. Fazio

## Abstract

Recent work has found that an individual’s beliefs and personal characteristics can impact perceptions of and responses to the COVID-19 pandemic. Certain individuals—such as those who are politically conservative, endorse conspiracy theories, or who believe the threat of COVID-19 to be exaggerated—are less likely to engage in such preventative behaviors as social distancing. The current research aims to address whether these individual difference variables not only affect people’s subjective and behavioral reactions to the pandemic, but also whether they actually impact individuals’ likelihood of contracting COVID-19. In the early months of the pandemic, U.S. participants responded to a variety of individual difference measures as well as questions specific to COVID-19 and the pandemic itself. Four months later, 2,120 of these participants responded with whether they had contracted COVID-19. Nearly all of our included individual difference measures significantly predicted whether a person reported believing they had contracted COVID-19 as well as whether they had actually tested positive for the virus in this four-month period. Additional analyses revealed that all of these relationships were primarily mediated by whether participants held accurate knowledge about COVID-19. These findings offer useful insights for developing more effective interventions aimed at slowing the spread of both COVID-19 and future diseases. Moreover, some findings offer critical tests of the validity of such theoretical frameworks as those concerning conspiratorial ideation and disgust sensitivity within a real-world context.

---

One year after the beginning of the COVID-19 pandemic, more than 100 million individuals worldwide had been infected while the total number of COVID-related deaths surpassed two million (WHO, 2021). The toll of the virus has been particularly pronounced in the United States, which has accounted for approximately one-quarter of these cases and deaths (WHO, 2021; CDC, 2021a).

Healthcare institutions across the country have been overwhelmed, with many regions’ intensive care units reporting being at full capacity at numerous points throughout the COVID-19 pandemic (HHS, 2021). While the recent arrival of multiple COVID-19 vaccines brings hope for curbing the virus, rollout of these vaccines has proved slower than anticipated (CDC 2021c; Smith, 2021).

Until a vaccine is widely available, the primary means by which to limit the spread of coronavirus is by following public health guidelines to socially distance, wear a mask while near others, wash one’s hands frequently, and avoid unnecessary trips outside of the home (CDC, 2021b; Matrajt & Leung, 2020; Chughtai, Seale, & Macintyre, 2020). Many recent studies have focused on the efficacy of these behaviors in preventing the spread of COVID-19. Studies on social distancing have found that communities with greater rates of distancing have lower virus transmission rates (Matrajt & Leung, 2020; Feng, Marchal, Sperry, & Yi, 2020). Providing convergent support for the effectiveness of distancing, other work has shown that individuals who personally distance more are less likely to contract the virus (Fazio et al., 2021a). Mask use has also proven effective at reducing the transmission of coronavirus, provided the masks are constructed in line with recommendations of the CDC or World Health Organization (Chughtai, Seale, & Macintyre, 2020). Similarly, the effectiveness of handwashing and hand sanitizing in reducing disease transmission has been well established for years (Aiello, Coulborn, Perez, & Larson, 2008; Burton et al., 2011; Rabie & Curtis, 2006)—effects that extend to the current coronavirus pandemic as well (Kratzel et al., 2020).

Despite the wealth of scientific evidence attesting to their efficacy, public health pleas to comply with these preventative behaviors have been met with a variety of responses (Fazio et al., 2021b; Rothgerber et al., 2020; Oosterhoff & Palmer, 2020). While some people reliably engage in these preventative behaviors, others overtly oppose the guidelines. Even as the number of COVID-19 cases and deaths has reached critical levels, the U.S. has witnessed continued protests condemning stay-at-home orders and mask mandates (BBC, 2020; CDC, 2021a).

Scientific research examining the psychological factors that shape a person’s response to the virus has identified several individual differences that predict less concern about the virus, including greater political conservatism, endorsement of conspiracy theories, and valuing material self-interest or individual freedom over public safety (Fazio et al., 2021b; Oosterhoff & Palmer, 2020; Romer & Jamieson, 2020). Although this research has been informative regarding the possible sources of variability in people’s responses to the virus, many questions remain. First, and perhaps most importantly, the overwhelming majority of past research has examined *self-reported* attitudes and behavior towards the pandemic. There are a number of factors that are likely to shape people’s responses to such self-report measures, such as the strong normative (and sometimes even legal) pressures to engage in preventative measures. It is therefore unlikely that these measures are true representations of people’s real-world behavior (see, e.g., Fazio et al., 2021a, for evidence supporting this argument with respect to social distancing behavior). Additionally, past research has used a variety of independent and dependent measures, as well as diverse operationalizations of attitudes and self-reported behavior toward the virus/pandemic, making comparisons across studies (e.g., of effect sizes) difficult. These factors further complicate the prospect of gaining an understanding of which factors are more or less predictive of responses to the COVID-19 pandemic.

In this research, we attempt to fill these theoretical and empirical gaps by going beyond self-reported attitudes and behaviors to understand the factors that prospectively predict *whether an individual actually contracts COVID-19 over time*. To our knowledge, this is the first study to assess the predictive power of individual differences in beliefs and personal characteristics with respect to the likelihood of contracting the virus. To address this question, we conducted a longitudinal study of *N* = 2,120 participants. At Time 1, shortly after the pandemic became a heightened concern for most Americans (Spring 2020), we assessed a wide variety of relevant beliefs, personality characteristics, and demographic factors (e.g., perceptions of the pandemic, trust in scientists, objective knowledge about COVID-19, political ideology, race/ethnicity). We then followed up with these same participants four months later to assess whether they had contracted the novel coronavirus in the intervening time period. Using these data, we examine which individual differences predict subsequent illness and which do not.

## Original Study

As highlighted previously, compliance with public health directives is necessary to curb the spread of COVID-19. Critically, however, the compliance literature lacks a general theoretical framework by which to explain who will comply with a given directive and why (Nezlek & Smith, 2017). To help fill this gap, in a recent study we (Fazio et al., 2021b) presented and tested a general theoretical framework of compliance, elucidating three key components that influence whether or not an individual will comply with any given behavioral directive: perceptions of the *source* of the directive, perceptions of the *challenge* that prompted the directive, and relevant *personal characteristics* of the target. The pandemic offered a unique opportunity to test this new framework within the context of COVID-19 and the accompanying directives to engage in preventative behaviors.

Regarding the first of these three broad components, the source(s) of the directive, this research identified public health officials and government officials as the two primary sources of the COVID-19 preventative behavior guidelines. As such, participants were asked to indicate their attitudes toward scientists, as perceptions of scientists were expected to directly impact one’s willingness to comply with science-based directives. Participants also reported trust in then-President Trump and the federal government as a whole. This distinction was made given that throughout the pandemic, government officials—including President Trump—have offered contradicting messages about the seriousness of COVID-19 and the need to engage in preventative behaviors (LeBlanc & Diamond, 2020; Siemaszko, 2020; Shepherd, 2020; Antonia Farzan, 2020; Paz, 2020).

The second component of the compliance framework is perceptions of the challenge itself—in this case, the COVID-19 pandemic and its surrounding context. The critical variables within this component were individuals’ perceptions of COVID-19’s seriousness, beliefs about the virus’s impact on society, and perceptions of the directives to engage in preventative behaviors (e.g., their efficacy). In addition to subjective perceptions of the pandemic, Fazio and colleagues (2021b) also included a measure of objective knowledge of the virus, anticipating that endorsing less correct information—or more *mis*information—about COVID-19 would predict less compliance with the directive.

The third and final component concerns relevant characteristics of the targets of the directive themselves that may influence receptivity. Perhaps unsurprisingly, given the salience, severity, and politicization of the pandemic, this research identified a wide range of relevant characteristics. First, the original study assessed participants’ personal sensitivity to disgust as well as their perceived vulnerability to disease in general (Fazio et al., 2021b). Since the pandemic has been widely politicized (Hart, Chinn, & Soroka, 2020), participants also reported their political orientation and which popular news sources they use. Conspiracy theories have also grown in popularity as is common during times of social distress and uncertainty (Brotherton, 2015; Uscinski & Parent, 2014). As such, participants reported endorsement of a variety of types of conspiracy theories, with the purpose being to assess whether greater endorsement of conspiracy theories might lead to less support for public health messaging.

Fazio et al. (2021b) focused specifically on the relationship between these various predictor variables and social distancing, using both self-reported and “virtual” (i.e., interactive graphical) social-distancing measures. As predicted, all of the previously discussed variables did in fact significantly relate to these measures of social distancing. For example, objective knowledge of COVID-19, disgust sensitivity, and trust in scientists were all positively associated with greater social distancing. Meanwhile, confidence in President Trump, conservatism, and greater endorsement of conspiracy theories negatively related to distancing behavior. Moreover, a later longitudinal study revealed that participants’ scores on the virtual measure of social distancing behavior were predictive of the likelihood of their subsequently contracting COVID-19 (Fazio et al., 2021a).

However, there are several reasons to question whether and to what degree each of these individual difference factors would translate into a higher likelihood of actually contracting the virus. For example, Fazio et al. (2021a) focused exclusively on social distancing behavior and did not examine any of the other various preventative behaviors that have been shown to reduce disease transmission, such as hand washing and mask-wearing. Further, several of these associations, although statistically robust, were quite small, and it may be that they are not sufficient to have real-world impacts on disease transmission. Thus, the critical question—whether and to what degree these various source, context, and target factors predict actual COVID-19 contraction—remains unanswered. We address this question in the present research.

## The Current Research

In the current study, we followed up with participants from the original research four months later to assess whether or not they contracted COVID-19. This allowed us to assess which beliefs, personality characteristics, and demographic factors prospectively predicted contracting the virus during the intervening period of time, as well as the relative magnitude of each of these effects. In doing so, we are also able to test critical theoretical questions regarding each of these diverse factors—many of which are theorized to predict illness. For example, past work predicts that individuals who are more sensitive to disgust will be less likely to contract a given illness, as disgust (at least in theory) motivates avoidance of potential pathogens in the environment (Tybur, Lieberman, Kurzban, DeScioli, 2013). Presumably, such avoidance also will promote preventative behaviors that decrease the likelihood of contracting illness. In addition, theoretical models of conspiratorial ideation suggest that people who are prone to conspiracy beliefs are likely to be relatively dismissive of anxiety-provoking events such as the current pandemic, as they adopt conspiracy theories as means by which to gain a sense of control over these kinds of situations (Douglas, Sutton, & Cichocka, 2017). Moreover, such conspiratorial tendencies are likely to promote both inattentiveness to true facts and a failure to reject misinformation about the event. As a result, individuals prone to conspiratorial ideation should be more likely to contract the virus. The current study offers an opportunity to test such predictions in a real-world context.

All data are available at https://osf.io/ywv5r/?view_only=6dd2b2715fe349bd8b2252624d25ad3d for researchers who wish to replicate or extend our analyses.

## Methods

We recruited our participant samples from Amazon Mechanical Turk (see Buhrmester, Kwang, & Gosling, 2011). Although not representative of the U.S. population, MTurk samples tend to be more demographically, politically, and geographically diverse than the samples typically used in psychological research (Paolacci & Chandler, 2014). They also perform similarly to non-MTurk samples across many tasks and measures (Berinsky, Huber, & Lenz, 2012; Hauser, Paolacci, & Chandler, 2019), including surveys on social and political attitudes (Clifford, Jewell, & Waggoner, 2015). Most importantly, however, our aim is not to make claims regarding the absolute frequency of COVID-19 illness in the population. Rather, we seek to understand which beliefs and characteristics predict whether an individual contracts the coronavirus over time. Given this aim, MTurk participants offer an appropriate test of our research questions.

Given the large number of variables we measured in these studies, we employed a “planned missing” design (Graham, 2012) in which different subgroups of participants completed different sets of predictor measures. First, all participants completed a set of measures regarding their beliefs and behaviors concerning the pandemic, as well as various demographic items and relevant covariate measures. We refer to this common set of measures as “the core survey.” Participants were then randomly assigned to one of four subsets of the remaining predictor variables, which were grouped according to theoretical and empirical relatedness. The four sub-studies included: (a) beliefs about the sources of the directive, (b) news sources and endorsement of conspiracy theories, (c) general interpersonal compassion, and (d) disgust sensitivity and perceived vulnerability to disease.

### Participants

The sample consisted of MTurk workers who had participated in one of two studies conducted in Spring 2020. All participants who had granted permission to re-contact them were invited to complete a brief survey approximately four months after their initial study for a payment of $1. A total of 2,120 individuals, all US residents, completed this survey (1,031 women, 1,074 men, 15 no response; *M*_age_ = 40.39, *SD*_*age*_ = 15.34). Study 1 was completed on May 7-8 (*n* = 1281)^1^ and Study 2 on June 9 (*n* = 839).

### Measures

After providing informed consent, participants completed a wide range of questions regarding the COVID-19 pandemic. These included the critical questions described above concerning their perceptions of the pandemic (e.g., whether it has been exaggerated; worry about contracting the virus), a test assessing knowledge about COVID-19, the subset of predictor variables to which they had been randomly assigned, and a series of demographic questions. In addition, participants also completed a set of measures concerning their behavior regarding the pandemic (e.g., self-reported behaviors, virtual measures of social distancing). Given that the relation between these latter measures of social distancing behavior and contracting COVID-19 were the focus of an earlier report (Fazio et al., 2021a), they are not analyzed here.

#### Perceptions of the pandemic

Participants completed a set of items regarding their perceptions of the pandemic. They were first asked how worried they were about personally contracting the novel coronavirus, how likely they thought they were to contract the virus, and how concerned they were about the spread of the virus in general. The last item asked whether they believed that the threat of COVID-19 had been “greatly exaggerated,” “somewhat exaggerated,” “adequately conveyed,” or “not conveyed strongly enough.”

#### COVID-19 knowledge

This brief test of objective COVID-19 knowledge consisted of 13 statements, all either facts or myths about COVID-19, based on information from the Centers for Disease Control and Prevention (CDC) and the World Health Organization (WHO). Participants were asked to indicate whether each statement was true or false. Examples of true statements include “Some individuals who have COVID-19 / the coronavirus do not show any symptoms” and “Washing one’s hands with soap and water for at least 20 seconds can reduce the spread of COVID-19 / the coronavirus.” Meanwhile, false statements included “Antibiotics are an effective treatment for COVID-19 / the coronavirus” and “Spraying chlorine on my body will protect me even if COVID-19 / the coronavirus has already entered my system.” We summed the number of correct responses to create an index of objective knowledge (α = .64). We also computed the correct number of responses for both true and false items separately to independently assess the effects of acceptance of true information versus rejection of falsehoods (α = .63 and α = .84., respectively).

#### Faith in government

Throughout the pandemic, different—and sometimes contradictory— information has been communicated by various government entities. We asked participants to provide their perceptions of a few of these major entities within four items, each with seven-point response scales ranging from “Not at all” to “Very much.” For example, participants were asked whether they “trust President Trump to lead us effectively through the COVID-19 crisis.” They also rated their general confidence in both President Trump (“Generally speaking, how much confidence do you have in President Trump?”) and the federal government (“Generally speaking, how confident are you that the federal government will address the nation’s problems effectively?”).

#### Trust in scientists

To assess participants’ trust in scientists, we used a shortened version (the 11 items with the highest corrected item-total correlation) of a measure developed by Nadelson et al. (2014). Using a five-point scale ranging from “Strongly disagree” to “Strongly agree,” participants rated statements including “We should trust the work of scientists” and “We cannot trust scientists because they are biased in their perspectives” (reverse coded). We averaged across these items to compute a composite rating (α = .81).

#### Science literacy

The Civic Scientific Literacy Scale (Miller, 1998) was used to assess participants’ general scientific knowledge. As a note, this measure was included in only one of the two initial studies (Study 1, *n* = 1281). Participants indicated whether they agreed or disagreed with 11 statements, including “The Earth goes around the Sun once each year” and “Electrons are smaller than atoms”. The number of correct responses served as an indicator of participants’ general scientific literacy (α = .58).

#### Endorsement of conspiracy theories

Participants’ tendency towards conspiratorial ideation was measured with the Generic Conspiracist Beliefs scale (Brotherton, French, & Pickering, 2013), which assesses endorsement of a variety of generic conspiracy theories. The 15 items of this scale include statements such as “Technology with mind-control capacities is used on people without their knowledge,” “The government permits or perpetrates acts of terrorism on its own soil, disguising its involvement,” and “Evidence of alien contact is being concealed from the public.” Participants rated their endorsement of each statement on a five-point scale ranging from “Definitely not true” to “Definitely true,” and the average rating across the 15 items was computed (α = .96).

#### News Sources

Participants were asked to select all of the news sources from which they had gotten their news in the past week from the following list: CNN, Fox News, MSNBC, NPR, national newspapers and magazines, social media, and ABC, CBS, or NBC News, as well as the option “do not follow the news.” Participants who selected at least one news source were then asked to indicate which of these sources they considered to be their primary news source. For our purposes, we were especially interested in exposure to Fox News as other work has shown that following Fox News relates strongly to attitudes toward the pandemic (Jurkowitz & Mitchell, 2020).

#### General interpersonal compassion

Fourteen items from a scale developed by Davis (1983) were used to assess participants’ general compassion for others. Statements included “I often have tender, concerned feelings for people less fortunate than me” and “Before criticizing somebody, I try to imagine how I would feel if I were in their place.” Participants responded to each item using a five-point scale ranging from “Does not describe me well” to “Describes me very well.” Ratings were averaged to compute an overall metric of interpersonal compassion (α = .87).

#### Disgust Sensitivity

We assessed participants’ sensitivity to disgust using the five-item contamination subscale of the Disgust Scale-Revised (Haidt, McCauley, & Rozin, 1994; Olatunji et al., 2007). Three items (e.g., “A friend offers you a piece of chocolate shaped like dog-doo”) were rated using a five-point scale ranging from “Not disgusting at all” to “Extremely disgusting,” while the other three (e.g., “I never let any part of my body touch the toilet seat in a public washroom”) were rated using a five-point scale ranging from “Strongly disagree” to “Strongly agree.” The overall score for disgust sensitivity was computed by averaging across the five items (α = .70).

#### Perceived vulnerability to disease

Duncan, Schaller, and Park’s (2009) 15-item scale was used to assess participants’ perceptions of their likelihood of contracting disease or illness in general. Statements included “If an illness is ‘going around’, I will get it” and “It does not make me anxious to be around sick people” (reverse-scored). Participants rated their agreement with each item on a five-point scale from “Strongly disagree” to “Strongly agree.” After recoding the reverse-scored items, the average rating across the 15 items was computed (α = .73).

#### Preexisting Conditions

Having preexisting health conditions was identified a priori as a likely predictor of contracting COVID-19. As such, participants were asked to consider their “personal health prior to the outbreak of the COVID-19 virus” and to then indicate whether they would have described themselves “as having pre-existing medical conditions that left you more vulnerable to the virus than the average person” by selecting one of five response options ranging from “Definitely not” to “Definitely yes.”

#### Demographics

Other work has shown that certain social groups in the U.S. tend to have higher rates of infection. This includes the elderly who are more likely to contract the virus, as well as to have more complications with the disease. Additionally, systemic and structural factors in US society have led racial and ethnic minorities to have higher infection, hospitalization, and death rates than non-Hispanic White Americans (CDC, 2020; The Atlantic, 2020). For example, the COVID-19 death rate of Black Americans is reported to be more than twice that of White Americans. Given this, participants were asked to report their age, gender, race and/or ethnicity. Additionally, participants indicated their political ideology on a seven-point scale ranging from “Extremely liberal” to “Extremely conservative.”

#### Follow-up survey

Four months after the initial study, participants completed a brief survey assessing whether they had or had not contracted the coronavirus. They were first asked whether they had been tested for COVID-19. If so, they indicated whether the test showed that they had COVID-19. If they had not been tested, they were asked “Even though you may not have been tested, do you believe that you have ever had COVID-19 / the coronavirus?” to which they responded yes or no. Participants who reported either testing positively or believing they had COVID-19 were then asked to select from a list of possibilities to indicate how they thought they might have contracted the virus.

## Results

### Descriptive Data

The follow-up survey first inquired as to whether participants had been tested for COVID-19. Of the 516 participants (24.3% of the total sample of 2,120) who reported having been tested, 116 (5.5% of the total sample) indicated that the test result was positive. Participants who had not been tested were asked whether they nevertheless believed they had contracted the coronavirus. Two-hundred and thirty-two participants (10.9% of the total sample) responded affirmatively. Thus, a total of 348 participants (16.42%) reported having experienced COVID-19 illness at the time of the follow-up survey.

### Predicting Reports of Having Contracted COVID-19 at Follow-up

Our initial analyses focused on the comparison of those participants who reported not having contracted COVID-19 (coded as 0) to those who reported either having tested positively or believing they had contracted COVID-19 despite having not been tested (coded as 1). Our major interest is to examine whether each of our variables of interest *prospectively predicts* subsequent illness. Hence, we excluded from the analyses any participants who reported having COVID-19 at the time of the initial study. Of the 2,120 follow-up participants, 235 had either reported a positive test result at Time 1 or reported that they believed they had COVID-19 but had not been tested. These participants were therefore excluded from analyses, resulting in a total sample of 1,885 participants for our first set of analyses. However, the sample size available for any given variable varies as a consequence of the planned missing design employed in the initial studies. That is, because some variables were randomly assigned only to specific subsamples of participants, our analyses for these variables are limited to the participants who completed those measures. Accordingly, in the tables that follow, we also include sample size details for each predictor variable.

To determine which individual difference factors predicted contracting COVID-19, we conducted a series of binary logistic regression analyses examining the dichotomous COVID-19 status variable at follow-up (i.e., did versus did not contract COVID-19) as a function of each of the predictor variables. This analysis allowed us not only to assess which variables predicted contracting COVID-19, but also the effect size for each prediction offered by the odds ratio—that is, how the odds of contracting COVID-19 change as a function of a unit change in the predictor variable. The results are summarized in Table 1, which presents, for each variable, the number of participants who did versus did not report having contracted the virus and the regression statistics. (To ease interpretation, all continuous predictor variables were standardized.) We highlight the key findings below.

**Table 1.** Predicting Reports of COVID-19 Status^a^.

|  | *n<sub>1</sub>/n<sub>2</sub>* | *B* | Wald | *p* | Odds Ratio |
| --- | --- | --- | --- | --- | --- |
| **Beliefs about the Source** |  |  |  |  |  |
| Trust in Scientists<sup>b</sup> | 59/452 | -.356 | 7.109 | .008 | .701 |
| Trust President Trump re COVID-19 crisis<sup>b</sup> | 59/451 | .359 | 7.334 | .007 | 1.432 |
| Confidence in Federal Gov't Effectiveness<sup>b</sup> | 59/451 | .409 | 8.534 | .003 | 1.506 |
| General Confidence in President Trump<sup>b</sup> | 59/451 | .282 | 4.503 | .034 | 1.326 |
| **Beliefs about the Context** |  |  |  |  |  |
| Worry about Contracting Virus<sup>b</sup> | 199/1686 | .409 | 28.121 | .000 | 1.506 |
| Likely to Contract Virus<sup>b</sup> | 199/1686 | .590 | 57.816 | .000 | 1.804 |
| Threat (not) exaggerated<sup>b</sup> | 199/1686 | -.430 | 35.573 | .000 | .651 |
| COVID Knowledge<sup>b</sup> | 199/1686 | -.680 | 141.664 | .000 | .507 |
| Acceptance of True Items<sup>b</sup> | 199/1686 | -.467 | 75.038 | .000 | .627 |
| Rejection of False Items<sup>b</sup> | 199/1686 | -.655 | 127.062 | .000 | .520 |
| **Other Receptivity-Related Characteristics** |  |  |  |  |  |
| General interpersonal compassion<sup>b</sup> | 49/487 | -.233 | 2.562 | .109 | .792 |
| Disgust Sensitivity<sup>b</sup> | 62/485 | .512 | 13.386 | .000 | 1.669 |
| Perceived vulnerability to disease<sup>b</sup> | 62/486 | .328 | 5.841 | .016 | 1.389 |
| Preexisting conditions<sup>b</sup> | 199/1684 | .350 | 25.443 | .000 | 1.420 |
| Political ideology (higher, more conservative)<sup>b</sup> | 198/1686 | .229 | 9.567 | .002 | 1.257 |
| Belief in conspiracy theories<sup>b</sup> | 59/464 | .664 | 20.425 | .000 | 1.943 |
| Science Literacy<sup>b</sup> | 29/249 | -.401 | 4.602 | .032 | .670 |
| Fox News<sup>c</sup> | 125/903 | .343 | 7.472 | .006 | 1.409 |
| NPR<sup>c</sup> | 125/903 | -.851 | 12.005 | .001 | .427 |
| Papers, Magazines<sup>c</sup> | 125/903 | -.363 | 7.436 | .006 | .695 |
| **Demographic Variables** |  |  |  |  |  |
| Age | 198/1686 | -.028 | 17.681 | .000 | .972 |
| Gender (1=male/0=female) | 198/1674 | .157 | 1.085 | .298 | 1.170 |
| Race (1=Black/0=White) | 175/1438 | .996 | 25.133 | .000 | 2.708 |

<sup>a</sup> coded 0 = No report of COVID-19 (*n<sub>2</sub>*), 1 = Tested Positively or Untested but believe had COVID-19 (*n<sub>1</sub>*)
<sup>b</sup> Standardized
<sup>c</sup> coded 0=neither watch last week, nor primary news source, 1=watched last week or primary, 2=both

We first examined our broad group of predictor variables that concerned perceptions of the source of the directive. Strikingly, all four of the source variables significantly prospectively predicted whether an individual contracted COVID-19. Greater trust in scientists, consistent with our predictions, was associated with reduced likelihood of contracting the virus. Conversely, more positive evaluations of President Trump were predictive of an increased likelihood of contracting the virus. Similarly, greater trust in the federal government also predicted a greater likelihood of contracting the virus, perhaps due to Trump’s strong association with the federal government (the correlation between each of the measures regarding Trump and that concerning the federal government was *r* = .71).

We next turned to the broad group of factors concerning people’s perceptions of the nature and context of the challenge itself. Here, too, we identified a number of predictors of whether an individual contracted the virus. In particular, the more that participants perceived that the threat posed by the pandemic had been exaggerated, the more likely they were to personally contract the coronavirus.

Interestingly, participants also displayed striking insight regarding their personal risks of contracting COVID-19. Those who expressed greater worry about contracting COVID-19, and those who thought they were more likely to contract it, were, in fact, more likely to actually contract the virus. Objective knowledge about the virus also seemed to play an important role in determining whether an individual contracted it: Having less knowledge about the virus predicted a greater likelihood of subsequently contracting COVID-19. This relation was evident both for the rejection of true statements and the endorsement of misinformation.

Finally, we turned to the third broad group of factors: relevant traits and characteristics of the targets themselves. Here, too, we identified several critical predictors of contracting the virus. Greater political conservatism (versus liberalism) was associated with a higher likelihood of contracting COVID-19. Having a greater general propensity towards conspiratorial ideation, too, was significantly associated with an increased likelihood of contracting the virus. Conversely, science literacy was negatively associated with illness, such that increased knowledge and understanding of scientific concepts predicted a lower likelihood of contracting COVID-19. Finally, an individual’s preferred news sources also appeared to have implications for contracting the virus, with three of the individual news sources being related to subsequent illness. Most notably, viewers of Fox News were more likely to contract COVID-19, whereas participants more heavily involved with NPR were less likely to do so.

Those who reported having pre-existing conditions that made them more vulnerable to the virus were considerably more likely to contract the virus during the subsequent four months. The same was true of participants who rated their general vulnerability to disease to be relatively high. Although the receptivity-related characteristics listed in Table 1 generally showed the expected relation with contracting COVID-19, there was one notable exception: The results for disgust sensitivity appear to directly contradict the dominant theoretical perspectives on the nature and function of disgust (e.g., Haidt, McCauley, & Rozin, 1994; Tybur et al., 2013). That is, past research and theory posit that greater disgust sensitivity serves to minimize an individual’s contact with potentially pathogenic substances, and thereby reduce one’s likelihood of illness. In direct contrast to these predictions, however, individuals who were more sensitive to disgust were actually more likely to contract COVID-19.

With respect to the demographic variables, age was a significant, albeit relatively weak factor. Younger age was associated with greater likelihood of contracting the virus. A substantial association was evident for a contrast regarding race. Black participants exhibited far greater odds of contracting COVID-19 than did White participants, consistent with past research (e.g., CDC, 2020; The Atlantic, 2020).

### Predicting Positive Test Results at Follow-up

We next conducted a set of parallel analyses with a second outcome variable: whether participants reported having actually tested positively for COVID-19. This variable offers a useful opportunity for validation of the results reported above. One might argue that individuals with certain beliefs or characteristics were for some reason more or less likely to interpret any ambiguous physical symptoms as an indication of having the virus. In contrast, testing positively for COVID-19 is considerably less ambiguous; it is a clear and salient event that seems relatively unlikely to be mis-construed or mis-reported.

Consistent with our previous analyses, the analyses for this variable excluded any participants who had reported a positive test result during their initial survey, allowing us to specifically test whether and how each of these predictors prospectively predicted a positive test result. The resulting sample involved 85 participants who subsequently tested positively (coded as 1) and 1,993 who reported either a negative test or not having been tested at all (coded as 0). Once again, however, these numbers apply only to the variables that had been included in the core survey completed by all participants at Time 1; sample sizes for each individual predictor differ as a function of the number of participants who completed that specific measure. Sample sizes and binary logistic regression statistics for each variable are presented in Table 2.

**Table 2.**
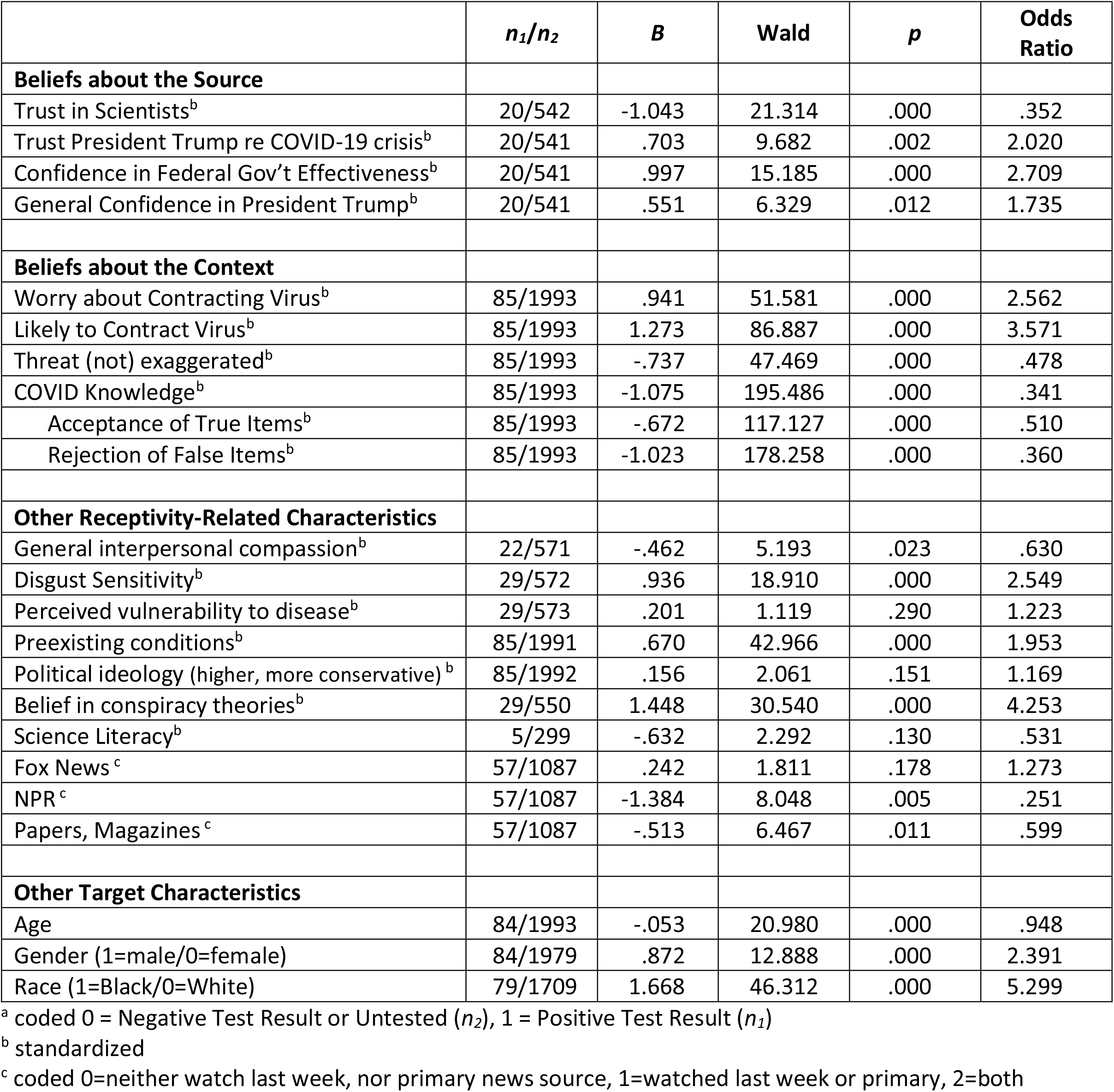
Predicting COVID-19 Positive Test^a^.

| | $n_1/n_2$ | $B$ | Wald | $p$ | Odds Ratio |
| --- | --- | --- | --- | --- | --- |
| **Beliefs about the Source** |  |  |  |  |  |
| Trust in Scientists<sup>b</sup> | 20/542 | -1.043 | 21.314 | .000 | .352 |
| Trust President Trump re COVID-19 crisis<sup>b</sup> | 20/541 | .703 | 9.682 | .002 | 2.020 |
| Confidence in Federal Gov't Effectiveness<sup>b</sup> | 20/541 | .997 | 15.185 | .000 | 2.709 |
| General Confidence in President Trump<sup>b</sup> | 20/541 | .551 | 6.329 | .012 | 1.735 |
| **Beliefs about the Context** |  |  |  |  |  |
| Worry about Contracting Virus<sup>b</sup> | 85/1993 | .941 | 51.581 | .000 | 2.562 |
| Likely to Contract Virus<sup>b</sup> | 85/1993 | 1.273 | 86.887 | .000 | 3.571 |
| Threat (not) exaggerated<sup>b</sup> | 85/1993 | -.737 | 47.469 | .000 | .478 |
| COVID Knowledge<sup>b</sup> | 85/1993 | -1.075 | 195.486 | .000 | .341 |
| Acceptance of True Items<sup>b</sup> | 85/1993 | -.672 | 117.127 | .000 | .510 |
| Rejection of False Items<sup>b</sup> | 85/1993 | -1.023 | 178.258 | .000 | .360 |
| **Other Receptivity-Related Characteristics** |  |  |  |  |  |
| General interpersonal compassion<sup>b</sup> | 22/571 | -.462 | 5.193 | .023 | .630 |
| Disgust Sensitivity<sup>b</sup> | 29/572 | .936 | 18.910 | .000 | 2.549 |
| Perceived vulnerability to disease<sup>b</sup> | 29/573 | .201 | 1.119 | .290 | 1.223 |
| Preexisting conditions<sup>b</sup> | 85/1991 | .670 | 42.966 | .000 | 1.953 |
| Political ideology (higher, more conservative)<sup>b</sup> | 85/1992 | .156 | 2.061 | .151 | 1.169 |
| Belief in conspiracy theories<sup>b</sup> | 29/550 | 1.448 | 30.540 | .000 | 4.253 |
| Science Literacy<sup>b</sup> | 5/299 | -.632 | 2.292 | .130 | .531 |
| Fox News<sup>c</sup> | 57/1087 | .242 | 1.811 | .178 | 1.273 |
| NPR<sup>c</sup> | 57/1087 | -1.384 | 8.048 | .005 | .251 |
| Papers, Magazines<sup>c</sup> | 57/1087 | -.513 | 6.467 | .011 | .599 |
| **Other Target Characteristics** |  |  |  |  |  |
| Age | 84/1993 | -.053 | 20.980 | .000 | .948 |
| Gender (1=male/0=female) | 84/1979 | .872 | 12.888 | .000 | 2.391 |
| Race (1=Black/0=White) | 79/1709 | 1.668 | 46.312 | .000 | 5.299 |

<sup>a</sup> coded 0 = Negative Test Result or Untested ( $n_2$ ), 1 = Positive Test Result ( $n_1$ )<sup>b</sup> standardized<sup>c</sup> coded 0=neither watch last week, nor primary news source, 1=watched last week or primary, 2=both

The results of these analyses largely concurred with those presented above involving the reports of having experienced COVID-19. Only a few variables for which a relation had been observed previously did not achieve statistical significance: perceived vulnerability to disease, political ideology, Fox News viewership, and science literacy. In contrast, while its earlier relation with illness did not reach a conventional level of significance, interpersonal compassion was statistically significant when predicting positive test results. The more participants described themselves as being generally compassionate, the less likely they were to have tested positively during the subsequent four months.

Indeed, a large number of the predictor variables that we examined were associated with positive test results. The effects of trust in scientists, belief that the threat posed by the pandemic had not been exaggerated, COVID-19 knowledge, and age were especially noteworthy in the extent to which they negatively predicted COVID-19 illness. Conversely, trust in former President Trump), confidence in the federal government, worry about contracting the virus, perceived likelihood of personally contracting the virus, disgust sensitivity, and conspiratorial ideation were associated with markedly increased odds of a positive COVID-19 test result during the subsequent four-month period. With respect to the demographic variables, younger participants, males, and Black (compared to White) individuals were more likely have had a positive test outcome.

### The Mediating Role of COVID-Specific Predictors

Next, we investigated potential means by which these various individual differences impact a person’s likelihood of contracting COVID-19. That is, we sought to gain insight into *why and how* these various beliefs, personality characteristics, and demographic factors influence a person’s likelihood of contracting the virus. Within the current study, predictors addressing one’s beliefs about the context would have developed as the pandemic unfolded (e.g., perceived severity of the threat, knowledge about COVID-19). Meanwhile, other variables we use to predict illness are largely individual differences which presumably would have characterized our participants prior to the pandemic (e.g., political ideology, disgust sensitivity, demographic factors). Consistent with the logic we laid out above, we anticipated that many of these pre-existing individual differences would influence disease contraction indirectly—via intermediary perceptions and beliefs about the pandemic—which, in turn, would lead a person to behave in ways that increase the probability they would contract the virus.

All our variables addressing perceptions of the pandemic proved to be strong predictors of testing positively for COVID-19 (see Table 2). These were COVID-19 knowledge, believing the threat of COVID-19 to (not) be exaggerated, the degree to which one is worried about contracting the virus, and the perceived likelihood of contracting the virus. Given that the latter two were strongly correlated (r= 70), each was standardized and then averaged together to form a composite measure we refer to as participants’ perceived risk of contracting COVID-19. Overall, these three variables directly concern people’s perceptions of the pandemic, and as such represent three potential paths by which a given individual difference might relate to one’s risk of contracting COVID-19. Specifically, we hypothesize that our individual difference measures are indirectly predicting illness by leading people to (a) develop less accurate (or more inaccurate) knowledge about COVID-19, (b) minimize the overall threat of the pandemic, and/or (c) accurately perceive that they have a greater risk of contracting the virus.

We performed a series of mediational analyses examining the relation between each of our individual difference variables and testing positive for COVID-19, mediated by each of the three COVID-specific variables. By considering the mediational variables simultaneously, we assess the unique variance attributable to each. That is, we assess the extent to which the relationship between a given individual difference and COVID-19 illness is uniquely mediated by accurate knowledge regarding COVID-19, assessment of the threat posed by the pandemic, and/or perceived risk of contracting the virus. Of course, mediation analyses cannot provide decisive evidence of causal process. Nevertheless, in identifying that factors that statistically account for the relations between our predictor variables and contracting the virus, they can provide some tentative insight into the process by which each of these factors affects probability of disease contraction. Figure 1 offers the standard structure for these mediational models.

**Figure 1.**
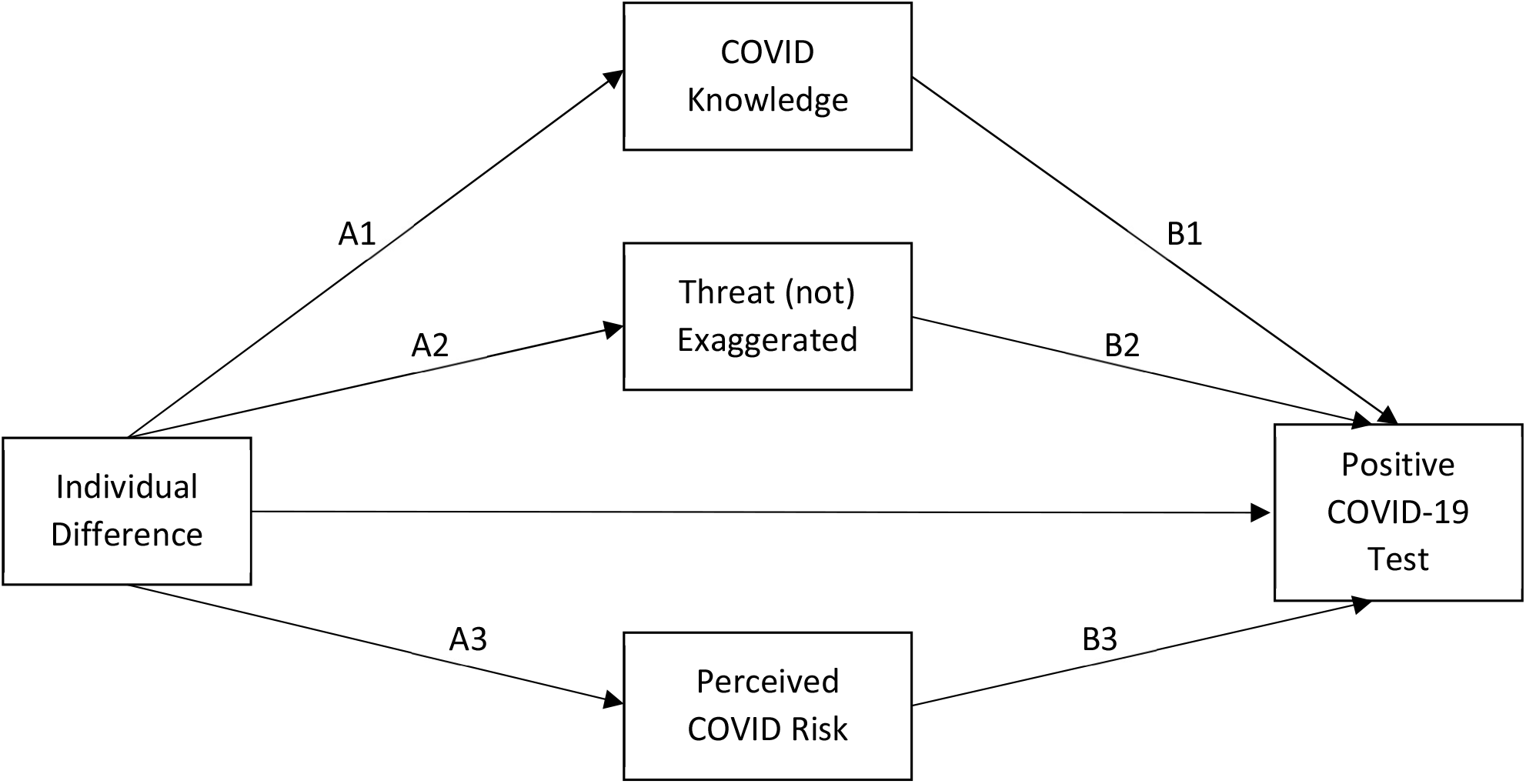
Basic Structure of Mediational Models Predicting Impact of Individual Difference Variables on Positive COVID-19 Tests via COVID-Specific Variables.

The results are summarized in Table 3, in which we present standardized beta coefficients for each individual difference variable as it relates to the COVID-specific variables (Figure 1 paths A1, A2, and A3). We also include estimates of the overall indirect effects of these individual differences on illness via each of the three mediators (Figure 1 paths A1*B1, A2*B2, and A3*B3). These estimates— which are themselves the products of the respective A and B paths (e.g., A1*B1)—allow us to assess the extent to which each of these COVID-specific variables accounts for the relationships we uncovered between our individual difference measures and COVID-19 illness. Full mediational models for each individual difference variable are included in the supplementary information.

**Table 3.** Indirect Effects of Individual Difference Variables on Testing Positive for COVID-19 via COVID-Specific Variables.

|  | COVID Knowledge |  | Threat (not) Exaggerated |  | Perceived COVID Risk |  |
| --- | --- | --- | --- | --- | --- | --- |
|  | Standardized Beta (A1) | Indirect effect (A1*B1) | Standardized Beta (A2) | Indirect effect (A2*B2) | Standardized Beta (A3) | Indirect effect (A3*B3) |
| **Beliefs about the Source** |  |  |  |  |  |  |
| Trust in Scientists | .581*** | -.379<sup>†</sup> | .552*** | -.116 | .059 | .058 |
| Trust President Trump re COVID-19 crisis | -.486*** | .413<sup>†</sup> | -.538*** | .173 | -.117** | -.112<sup>†</sup> |
| Confidence in Federal Gov't Effectiveness | -.367*** | .236<sup>†</sup> | -.340*** | .061 | -.059 | -.058 |
| General Confidence in President Trump | -.460*** | .410<sup>†</sup> | -.536*** | .206 | -.128** | -.126<sup>†</sup> |
| **Other Receptivity-Related Characteristics** |  |  |  |  |  |  |
| General interpersonal compassion | .196*** | -.163<sup>†</sup> | .219*** | -.015 | .074 | .013 |
| Disgust Sensitivity | -.320*** | .306<sup>†</sup> | .082 | .001 | .271*** | .321<sup>†</sup> |
| Perceived vulnerability to disease | .030 | -.028 | .196*** | .002 | .419*** | .464<sup>†</sup> |
| Preexisting conditions | -.203*** | .165<sup>†</sup> | .051* | -.010 | .364*** | .287<sup>†</sup> |
| Political ideology (higher, more conservative) | -.263*** | .224<sup>†</sup> | -.413*** | .087 | -.135*** | -.106<sup>†</sup> |
| Belief in conspiracy theories | -.509*** | .353<sup>†</sup> | .265*** | .127 | .102* | .102<sup>†</sup> |
| Science Literacy<sup>a</sup> | .434*** | -.310 | -.159** | -.073 | -.121* | -.212 |
| Fox News<sup>b</sup> | -.273*** | .284<sup>†</sup> | -.364*** | .114 | -.030 | -.028 |
| NPR<sup>b</sup> | .352*** | -.348<sup>†</sup> | .350*** | -.106 | .138** | .134<sup>†</sup> |
| Papers, Magazines<sup>b</sup> | .213*** | -.218<sup>†</sup> | .210*** | -.061 | .089* | .083<sup>†</sup> |
| **Other Target Characteristics** |  |  |  |  |  |  |
| Age | .073*** | -.051<sup>†</sup> | -.006 | .002 | -.025 | -.022 |
| Gender (1=male/0=female) | -.246*** | .198<sup>†</sup> | -.207*** | .036 | -.125** | -.102<sup>†</sup> |
| Race (1=Black/0=White) | -.1.052*** | .916<sup>†</sup> | .069 | .010 | .355*** | .247<sup>†</sup> |

<sup>a</sup> Statistics for this variable should be interpreted cautiously as the bootstrapped mediational model failed to converge, presumably due to the small number of positive test cases (5 out of 304).
<sup>b</sup> coded 0=neither watch last week, nor primary news source, 1=watched last week or primary, 2=both
\* $p < .05$ ; \*\* $p < .01$ ; \*\*\* $p < .001$
<sup>†</sup> denotes a 95% confidence interval that does not include zero

Interestingly, several of our individual difference measures differentially relate to the COVID-specific variables. When the three COVID-specific variables are entered together as potential mediators for the relationships between each of the four source variables and COVID illness, all four relationships appear to be mediated by COVID knowledge. This suggests that positive assessments of Trump and the federal government may have led people to develop less knowledge about the virus, while trust in science led people to develop greater knowledge. These knowledge differences, in turn, appear as though they may have prompted people to act in ways that affected their probability of contracting the virus.

The two variables specific to Trump (trust in Trump to lead us through the pandemic, and general confidence in Trump) were additionally mediated to a lesser degree by individuals’ insights regarding their perceived risk of contracting the virus. Interestingly, trust in Trump was associated with lower perceptions of one’s personal risk of contracting the virus. Although the direct relationship between these Trump-relevant variables and COVID illness is positive (i.e., more trust or confidence in Trump predicts greater likelihood of illness), the overall indirect effect of these Trump-relevant variables on illness via perceived risk of COVID-19 is negative. That is, greater confidence in Trump was associated with a decrease in perceived risk of COVID-19, and such individuals were in fact less likely to contract the virus.

Turning to the receptivity-related characteristics, we find that COVID knowledge acted as strong mediator for nearly all the variables. The relation between general compassion and illness was mediated by the development of more accurate knowledge, whereas the relations for disgust sensitivity, political conservatism, and conspiratorial ideation were mediated by less accurate knowledge. The relations with the various news sources also were mediated largely by the COVID knowledge variable, with Fox News being associated with less accuracy, and NPR and national newspapers and magazines with more accurate COVID information.

Insight into one’s perceived risk of contracting COVID-19 emerged as the primary mediator for the relations involving perceived vulnerability to disease and pre-existing conditions. Some of the other receptivity variables were also mediated by perceived risk, although generally to a lesser extent than the mediational role of COVID knowledge. Greater disgust sensitivity and greater conspiratorial ideation involved an accurate sense that one was more at risk, whereas more conservative individuals tended to perceive themselves to be at a lower risk of contracting COVID-19 (parallel to the findings for the Trump-relevant source variables). Finally, followers of NPR and national newspapers/magazines were also more likely to consider themselves at a higher risk of contracting COVID-19, and this insight proves correct in that they were *more* likely to contract the virus.

Lastly, we find that the relations of age, gender, and the Black/White race variable with testing positively for COVID-19 were all mediated primarily by their associations with the development of accurate knowledge. Younger individuals, males, and Black participants were likely to have less accurate knowledge about COVID-19 and were more likely to contract the virus. Gender and the race variable were also mediated, to a lesser extent, by perceptions of risk. Female and Black participants who more accurately perceive themselves to be at a higher risk are indeed more likely to have contracted COVID-19.

In summary, when accounting for potential mediating effects of the three COVID-specific variables simultaneously, COVID knowledge emerged as the primary mediator for the effects of our individual differences on COVID illness. Furthermore, we find that while additional variance in several of our mediational models was accounted for by participants’ assessments of their personal risk of contracting COVID-19, perceptions of the threat posed by the pandemic did not show a mediational effect for any of our predictors. In general, the more dominant pathway to COVID-19 illness appears to be the extent of accurate knowledge regarding the virus.

## Discussion

This research demonstrates the importance of an individual’s beliefs and personal characteristics for predicting whether said individual is likely to contract COVID-19. We identified several powerful predictors of contracting illness, including trust in the major sources of information about COVID-19 (e.g., scientists, the president), beliefs about the severity of the pandemic itself, personal insights about contracting the virus, and accurate knowledge about COVID-19. In addition, a number of other theoretically relevant individual characteristics were predictive of subsequent illness. Especially noteworthy were the associations involving conspiratorial ideation—which supported dominant theoretical models—and disgust sensitivity—which contradicted past research and theory. Thus, in addition to the immediate practical implications of these findings, the present research also provided critical tests of theoretical frameworks surrounding these individual differences in the important real-world context offered by the COVID-19 pandemic.

More generally, the findings offer clear evidence of a relation between individuals’ beliefs and a critically important health outcome. As such, these findings could be leveraged to reduce the spread of the COVID-19 virus. For example, many of our findings point to the politicized nature of the pandemic in the U.S. As many conservatives—including former President Trump—largely dismissed the significance of COVID-19 and/or occasionally conveyed misinformation about the virus, individuals on the political right are less likely to report engaging in preventative measures such as social distancing (Fazio et al., 2021b; Rothgerber et al., 2020; Grossman, Kim, Rexer, & Thirumurthy, 2020). Our research demonstrates that one’s political beliefs not only affect self-reported attitudes or beliefs, but one’s actual health. Thus, it is imperative that government and public health officials aim to reduce this political divide and avoid politicization of this and any future pandemics. Our data suggests a few paths by which to do so: by promoting the dissemination of accurate knowledge, dispelling misinformation, and undercutting conspiracy theories.

This work also revealed the significance of conspiratorial ideation. As noted earlier, belief in conspiracy theories has been shown to relate to a number of important political and health outcomes (Douglas, Sutton, & Cichocka, 2017; Lewandowsky, Gignac, & Oberauer, 2013). However, the present findings illustrate the significance of conspiratorial ideation in especially notable ways. Namely, the measure involved not any conspiracy theories that directly concerned COVID-19, but generic beliefs regarding such matters as alien contact and powerful, secretive forces (Brotherton et al., 2013). Past work has found systematic increases in conspiracy beliefs during times of turmoil, with some positing that conspiracy theories seem attractive during these times as they offer explanations that allow individuals to maintain a particular worldview (Brotherton, 2015; Uscinski & Parent, 2014). This explanation argues that conspiracy theories may act as a sort of buffer against stress or uncertainty. In doing so, though, these individuals may come to not only downplay the significance of the existing threat, but also develop less accurate knowledge of the threat. Our findings appear to fit this theoretical explanation, as conspiratorial ideation was associated strongly with less accurate knowledge regarding COVID-19—a factor which, in turn, increased the likelihood of contracting COVID-19. To our knowledge, this is the first demonstration that conspiracy beliefs prospectively predict a negative health outcome, while providing critical information regarding how these effects unfold.

Also of particular interest are the findings related disgust sensitivity, which appeared to run directly counter to theoretical models of disgust. Greater disgust sensitivity was associated not with a reduced likelihood of contracting the virus, but rather with an increased likelihood of doing so. This was true both for the reports of having gotten the virus and for testing positively. Hence, it is difficult to dismiss this finding as stemming from, e.g., more disgust-sensitive participants’ having possibly misconstrued a set of ambiguous symptoms. The direction of the relation poses a serious challenge to theoretical frameworks that view disgust sensitivity as a disease-avoidance mechanism (Oaten, Stevenson, & Case, 2009; Schaller, 2006; Schaller & Park, 2011). Our mediational analyses showed that the positive relationship between disgust sensitivity and testing positive for COVID-19 was mediated by both COVID knowledge and perceived COVID-19 risk, with disgust sensitivity being negatively related to COVID knowledge and positively related to assessments of personal risk. The latter suggests that the positive relation may reflect a propensity for individuals characterized by greater disgust sensitivity to be more insightful regarding their actual risk of contracting the virus, just as do those who report preexisting conditions or general vulnerability to disease.

An additional possibility is suggested by the observed mediational role of COVID-19 knowledge. Greater disgust sensitivity was associated with less accurate knowledge, which was itself predictive of a greater likelihood of testing positively for the virus. It may be that individuals characterized by more disgust sensitivity tended to avoid and/or not fully process the validity of information regarding the virus, possibly because they experienced disgust reactions in response to such information. Given the critical role knowledge appears to play, any such avoidance may have countered the postulated disease avoidance function of disgust sensitivity.

To further evaluate this possibility with the current data, we ran a binary logistic regression model with both disgust sensitivity and COVID-19 knowledge as predictors of illness. Within this model, COVID-19 knowledge was a significant predictor for both subjective reports of illness (*β* = –.73, *SE* = .11, Wald = 41.89, *p* < .001) and reports of having tested positively (*β* = –1.06, *SE* = .15, Wald = 50.40, *p* < .001), whereas disgust sensitivity was not (*β* = .19, *SE* = .16, Wald = 1.45, *p* = .228; *β* = .36, *SE* = .26, Wald = 1.95, *p* = .162). In other words, COVID-19 knowledge appears to attenuate the unexpected effect of disgust sensitivity on likelihood of contracting the virus. Future research will need to further address why disgust sensitivity did not have the postulated beneficial effects on avoiding COVID-19, and whether this calls into question the general theoretical premise regarding disgust sensitivity and disease avoidance, or whether the absence of any signs of a positive relation might be specific to other features of the COVID-19 pandemic and the surrounding context.

Finally, our last set of analyses (Table 3) aimed to further understanding of the relationships between our individual difference variables and COVID illness by examining the potential mediational roles of variables specific to the pandemic: COVID knowledge, assessments of the threat posed by the pandemic, and personal risk of contracting COVID-19. While we saw in the initial analyses (Tables 1 and 2) that accurate knowledge about COVID-19 was a strong predictor of illness, our mediational analyses reveal it to be a mechanism by which many pre-existing individual differences come to impact likelihood of contracting COVID-19. In fact, when controlling for the potential mediational effects of other COVID-specific variables (i.e., perception of the threat of the pandemic, personal insight about COVID-19), COVID knowledge was identified as the primary pathway relating our individual differences to the likelihood of testing positive for COVID-19. These mediational analyses suggest that increasing accurate COVID knowledge—and decreasing endorsement of misinformation—should reduce the negative impact of individual differences such as conspiratorial ideation, lack of trust in scientists, and political conservatism on likelihood of contracting the virus. As such, COVID knowledge could be a critical target for interventions aimed at decreasing the spread of COVID-19.

What is not yet known is exactly why certain individuals develop greater COVID knowledge. For example, the relationship between trust in scientists and testing positive for COVID-19 was mediated by COVID-19 knowledge in that those who trust scientists had higher rates of accurate knowledge, which decreased their likelihood of illness. It could be that individuals who are inclined to trust scientists purposely sought out more accurate information, that they tend to be in environments with better access to scientifically rigorous information, and/or that they approached misinformation with greater skepticism. Whatever the specific means, past work has shown that individuals with higher rates of knowledge are more likely to follow preventative measures (Fazio et al., 2021b; Zhou, Long, Kong, & Campy, 2020), which in turn makes them less likely to contract COVID-19 (Fazio et al., 2021a; Chughtai, Seale, & Macintyre, 2020; Fisher et al., 2020). While our data offers some unique support for this theoretical reasoning, future research would need to evaluate in a progressive, longitudinal manner the full path from individual difference variables to knowledge acquisition to voluntary preventative behaviors and, ultimately, illness.

In conclusion, the current research offers novel evidence regarding the importance of one’s beliefs and personal characteristics in predicting the likelihood of contracting the COVID-19 virus. The findings identify personal, psychological factors that appear to predispose a person to undue risk of contracting COVID-19. While some of the variables we assessed directly concerned the pandemic (e.g., knowledge about COVID-19, believing the threat of COVID-19 to be exaggerated), many were individual differences that were likely to have characterized the participants prior to the pandemic’s emergence (e.g., conspiratorial ideation, political ideology, interpersonal compassion). Yet, these too had meaningful effects. Moreover, our mediational analyses in particular provide initial evidence as to how and why each of these individual difference factors relate to disease contraction, offering critical insight for key points of intervention aimed at reducing the spread of COVID-19. We hope this work will inform future public health efforts to reduce transmission of not only the COVID-19 virus, but any future disease outbreaks as well.

### Context

Since the start of the pandemic, epidemiologists, public health professionals, politicians, and social scientists have attempted to address the devastating spread of COVID-19. While research has consistently demonstrated the effectiveness of preventative health behaviors like social distancing and mask use, many U.S. residents have consistently rejected directives to engage in these lifesaving behaviors. While some recent research has evaluated how certain personal characteristics relate to (self-reported) engagement in preventative behaviors, we still do not know whether—and which— specific individual differences predict contraction of COVID-19. In identifying numerous characteristics that alter a person’s risk of contracting the virus, this work provides unique insights into how public health officials, politicians, health psychologists, and other social scientists might develop more effective messaging and policies—as well as which groups and individuals should be most actively targeted by these campaigns—to curb the transmission of COVID-19 and similar diseases in the future.

## Supporting information

Supplemental mediational analyses

## Data Availability

Data has been made publicly available on the Open Science Framework.

https://osf.io/ywv5r/?view_only=6dd2b2715fe349bd8b2252624d25ad3d

## Footnotes

1 See Fazio, Ruisch, Moore, Granados Samayoa, Boggs, & Ladanyi (2021b) for a detailed report of Study 1.

## References

Aiello, A. E., Coulborn, R. M., Perez, V., & Larson, E. L. (2008). Effect of hand hygiene on infectious disease risk in the community setting: a meta-analysis. American journal of public health, 98(8), 1372–1381. https://doi.org/10.2105/AJPH.2007.124610

Antonia Farzan, R. (2020, October 09). WHO calls for ‘consistent messaging’ after Trump and Meadows contradict CDC chief. The Washington Post. https://www.washingtonpost.com/nation/2020/09/17/coronavirus-covid-live-updates-us/

BBC. (2020, April 21). Coronavirus lockdown protest: What’s behind the US demonstrations? News: US and Canada. https://www.bbc.com/news/world-us-canada-52359100

Berinsky, A. J., Huber, G. A., & Lenz, G. S. (2012). Evaluating online labor markets for experimental research: Amazon. com’s Mechanical Turk. Political analysis, 20(3), 351–368.

Brotherton, R. (2015). Suspicious minds: Why we believe conspiracy theories. Bloomsbury Publishing.

Brotherton, R., French, C. C. and Pickering, A. D. (2013). Measuring belief in conspiracy theories: The Generic Conspiracist Beliefs Scale. Frontiers in Psychology, 4: 279, 1–15.

Buhrmester, M., Kwang, T., & Gosling, S. D. (2016). Amazon’s Mechanical Turk: A new source of inexpensive, yet high-quality data?.

Burton, M., Cobb, E., Donachie, P., Judah, G., Curtis, V., & Schmidt, W. P. (2011). The effect of handwashing with water or soap on bacterial contamination of hands. International journal of environmental research and public health, 8(1), 97–104. https://doi.org/10.3390/ijerph8010097

Centers for Disease Control and Prevention (CDC; 2020, November 30). COVID-19 Hospitalization and Death by Race/Ethnicity. Cases, Data & Surveillance. https://www.cdc.gov/coronavirus/2019-ncov/covid-data/investigations-discovery/hospitalization-death-by-race-ethnicity.html

Centers for Disease Control and Prevention (CDC; 2021a, February 4). Trends in Number of COVID-19 Cases and Deaths in the US Reported to CDC, by State/Territory. CDC COVID Data Tracker. https://covid.cdc.gov/covid-data-tracker

Centers for Disease Control and Prevention (CDC; 20210b, February 4). How to Protect Yourself & Others. Your Health. https://www.cdc.gov/coronavirus/2019-ncov/prevent-getting-sick/prevention.html

Centers for Disease Control and Prevention (CDC; 2021c, January 5). 8 things to know about the U.S. COVID-19 vaccination program. https://www.cdc.gov/coronavirus/2019-ncov/vaccines/8-things.html

Chughtai, A. A., Seale, H., & Macintyre, C. R. (2020). Effectiveness of cloth masks for protection against severe acute respiratory syndrome coronavirus 2. Emerging infectious diseases, 26(10).

Clifford, S., Jewell, R. M., & Waggoner, P. D. (2015). Are samples drawn from Mechanical Turk valid for research on political ideology?. Research & Politics, 2(4), 2053168015622072.

Davis, M. H. (1983). The effects of dispositional empathy on emotional reactions and helping: A multidimensional approach. Journal of Personality, 51(2), 167–184.

Douglas, K. M., Sutton, R. M., & Cichocka, A. (2017). The psychology of conspiracy theories. Current Directions in Psychological Science, 26(6), 538–542.

Duncan, L. A., Schaller, M., & Park, J. H. (2009). Perceived vulnerability to disease: Development and validation of a 15-item self-report instrument. Personality and Individual differences, 47(6), 541–546.

Fazio, R. H., Ruisch, B. C., Moore, C. A., Samayoa, J. A. G., Boggs, S. T., & Ladanyi, J. T. (2021a). Social distancing decreases an individual’s likelihood of contracting COVID-19. Proceedings of the National Academy of Sciences, 118(8).

Fazio, R. H., Ruisch, B. C., Moore, C. A., Samayoa, J. A. G., Boggs, S. T., & Ladanyi, J. T. (2021). Who is (not) complying with the US social distancing directive and why? Testing a general framework of compliance with virtual measures of social distancing. PloS one, 16(2), e0247520.

Feng, Y., Marchal, T., Sperry, T., & Yi, H. (2020). Influence of wind and relative humidity on the social distancing effectiveness to prevent COVID-19 airborne transmission: A numerical study. Journal of aerosol science, 105585.

Fisher, K. A., Tenforde, M. W., Feldstein, L. R., Lindsell, C. J., Shapiro, N. I., Files, D. C., … & IVY Network Investigators. (2020). Community and close contact exposures associated with COVID-19 among symptomatic adults≥ 18 years in 11 outpatient health care facilities—United States, July 2020. Morbidity and Mortality Weekly Report, 69(36), 1258.

Graham, J. W. (2012). Missing data: Analysis and design. Springer Science & Business Media.

Grossman, G., Kim, S., Rexer, J. M., & Thirumurthy, H. (2020). Political partisanship influences behavioral responses to governors’ recommendations for COVID-19 prevention in the United States. Proceedings of the National Academy of Sciences, 117(39), 24144–24153.

Haidt, J., McCauley, C., & Rozin, P. (1994). Individual differences in sensitivity to disgust: A scale sampling seven domains of disgust elicitors. Personality and Individual differences, 16(5), 701–713.

Hart, P. S., Chinn, S., & Soroka, S. (2020). Politicization and Polarization in COVID-19 News Coverage. Science Communication, 1075547020950735.

Hauser, D., Paolacci, G., & Chandler, J. (2019). Evidence and Solutions. Handbook of Research Methods in Consumer Psychology, 319.

Jurkowitz, M., & Mitchell, A. (2020). Cable TV and COVID-19: How Americans perceive the outbreak and view media coverage differ by main news source. Pew Research Center.

Kratzel, A., Todt, D., V’kovski, P., Steiner, S., Gultom, M., Thao, T. T. N., … & Pfaender, S. (2020). Inactivation of severe acute respiratory syndrome coronavirus 2 by WHO-recommended hand rub formulations and alcohols. Emerging infectious diseases, 26(7), 1592.

LeBlanc, P., & Diamond, J. (2020, November 16). Trump coronavirus adviser Scott Atlas urges Michigan to ‘rise up’ against new Covid-19 measures. CNN Politics. https://www.cnn.com/2020/11/15/politics/scott-atlas-coronavirus-michigan/index.html

Lewandowsky, S., Gignac, G. E., & Oberauer, K. (2013). The role of conspiracist ideation and worldviews in predicting rejection of science. PloS one, 8(10), e75637.

Matrajt, L., & Leung, T. (2020). Evaluating the effectiveness of social distancing interventions to delay or flatten the epidemic curve of coronavirus disease. Emerging infectious diseases, 26(8), 1740.

Miller, J. D. (1998). The measurement of civic scientific literacy. Public understanding of science, 7, 203–223.

Nadelson, L., Jorcyk, C., Yang, D., Jarratt Smith, M., Matson, S., Cornell, K., & Husting, V. (2014). I just don’t trust them: the development and validation of an assessment instrument to measure trust in science and scientists. School Science and Mathematics, 114(2), 76–86.

Nezlek, J. B., & Smith, C. V. (2017). 4 CHAPTER Social Influence and Personality. The Oxford handbook of social influence, 53.

Oaten, M., Stevenson, R. J., & Case, T. I. (2009). Disgust as a disease-avoidance mechanism. Psychological bulletin, 135(2), 303.

Olatunji, B. O., Williams, N. L., Tolin, D. F., Abramowitz, J. S., Sawchuk, C. N., Lohr, J. M., & Elwood, L. S. (2007). The Disgust Scale: item analysis, factor structure, and suggestions for refinement. Psychological assessment, 19(3), 281.

Oosterhoff, B., & Palmer, C. A. (2020). Attitudes and psychological factors associated with news monitoring, social distancing, disinfecting, and hoarding behaviors among US adolescents during the coronavirus disease 2019 pandemic. JAMA pediatrics.

Paolacci, G., & Chandler, J. (2014). Inside the Turk: Understanding Mechanical Turk as a participant pool. Current directions in psychological science, 23(3), 184–188.

Paz, C. (2020, November 02). All the President’s lies about the Coronavirus. https://www.theatlantic.com/politics/archive/2020/11/trumps-lies-about-coronavirus/608647/

Rabie, T., & Curtis, V. (2006). Handwashing and risk of respiratory infections: a quantitative systematic review. Tropical medicine & international health : TM & IH, 11(3), 258–267. https://doi.org/10.1111/j.1365-3156.2006.01568.x

Romer, D., & Jamieson, K. H. (2020). Conspiracy theories as barriers to controlling the spread of COVID- 19 in the US. Social Science & Medicine, 263, 113356.

Rothgerber, H., Wilson, T., Whaley, D., Rosenfeld, D. L., Humphrey, M., Moore, A. L., & Bihl, A. (2020, April 22). Politicizing the COVID-19 Pandemic: Ideological Differences in Adherence to Social Distancing. https://doi.org/10.31234/osf.io/k23cv

Schaller, M. (2006). Parasites, behavioral defenses, and the social psychological mechanisms through which cultures are evoked. Psychological Inquiry, 17(2), 96–101.

Schaller, M., & Park, J. H. (2011). The behavioral immune system (and why it matters). Current Directions in Psychological Science, 20(2), 99–103.

Shepherd, K. (2020, November 16). Trump coronavirus adviser tells Michigan to ‘rise up’ against new shutdown orders. The Washington Post. https://www.washingtonpost.com/nation/2020/11/16/michigan-scott-atlas-coronavirus/

Siemaszko, C. (2020, October 07). Dr. Fauci contradicts Trump’s false claim that Covid-19 is as deadly as flu. NBC News. https://www.nbcnews.com/news/us-news/dr-fauci-contradicts-trump-s-false-claim-covid-19-deadly-n1242340

Smith, A. (2021, January 31). Covid vaccines: Rollout in disarray in U.S. and abroad. Retrieved from https://www.nbcnews.com/news/world/covid-vaccines-rollout-disarray-u-s-abroad-n1256144

The Atlantic. (2020). COVID-19 is affecting Black, Indigenous, Latinx, and other people of color the most. The COVID Racial Data Tracker. https://covidtracking.com/race

Tybur, J. M., Lieberman, D., Kurzban, R., & DeScioli, P. (2013). Disgust: evolved function and structure. Psychological review, 120(1), 65.

U.S. Department of Health and Human Services (HHS). Estimated ICU beds occupied by State Timeseries - COVID-19 ESTIMATED PATIENT impact and hospital capacity by state. (2021, February 1). https://healthdata.gov/dataset/covid-19-estimated-patient-impact-and-hospital-capacity-state/resource/82e733c6-7baa-4c65

Uscinski, J. E., & Parent, J. M. (2014). American conspiracy theories. Oxford University Press.

World Health Organization (WHO). WHO Coronavirus Disease (COVID-19) Dashboard. (2021, February 4). https://covid19.who.int/

Zhou, M., Long, P., Kong, N., & Campy, K. S. (2020). Characterizing Wuhan residents’ mask-wearing intention at early stages of the COVID-19 pandemic. Patient Education and Counseling.

