## Supplemental mediational analyses for "Contracting COVID-19: A Longitudinal Investigation of the Impact of Beliefs and Knowledge"

**Supplemental Materials**

Included here are all relevant statistics for our mediational models (reported partially in Table 3 of the main text). Each figure depicts the model in full with statistics included for all direct paths. Each table includes statistics for indirect effects within the model.

*Supplementary Figure 1. Direct effects of trust in scientists on COVID-19 test results mediated by COVID-specific predictors*

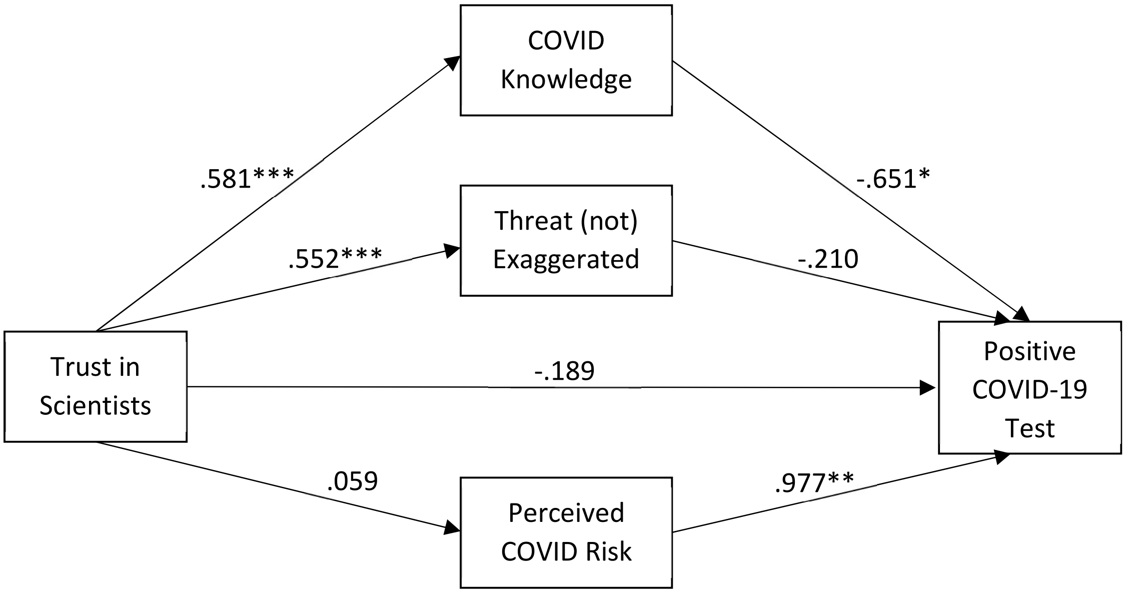


*$p < .05$; **$p < .01$; ***$p < .001$

*Supplementary Table 1. Indirect effects of trust in scientists on COVID-19 test results*

| | Indirect Effect | SE | 95% Confidence Interval |
|---|---|---|---|
| TOTAL | -.437† | .197 | [-.886, -.104] |
| COVID Knowledge | -.379† | .206 | [-.855, -.036] |
| Threat (not) Exaggerated | -.116 | .205 | [-.506, .286] |
| Perceived COVID Risk | .058 | .062 | [-.031, .206] |

†denotes a 95% confidence interval that does not include zero

*Supplementary Figure 2. Direct effects of trust in Trump (re COVID-19 crisis) on COVID-19 test results mediated by COVID-specific predictors*

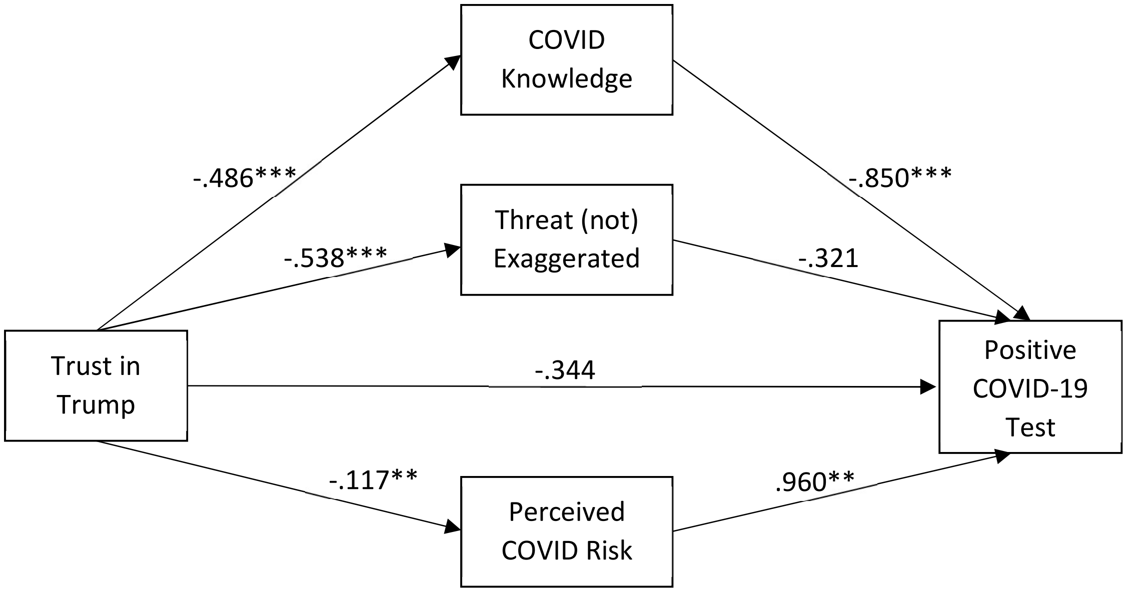


*$p < .05$; **$p < .01$; ***$p < .001$

*Supplementary Table 2. Indirect effects of trust in Trump (re COVID-19 crisis) on COVID-19 test results*

| | Indirect Effect | SE | 95% Confidence Interval |
|---|---|---|---|
| TOTAL | -.474$^{\dagger}$ | .219 | [.062, .937] |
| COVID Knowledge | .413$^{\dagger}$ | .157 | [.163, .774] |
| Threat (not) Exaggerated | .173 | .229 | [-.279, .647] |
| Perceived COVID Risk | -.112$^{\dagger}$ | .071 | [-.292, -.017] |

$^{\dagger}$denotes a 95% confidence interval that does not include zero

*Supplementary Figure 3. Direct effects of confidence in federal government (re COVID-19 crisis) on COVID-19 test results mediated by COVID-specific predictors*

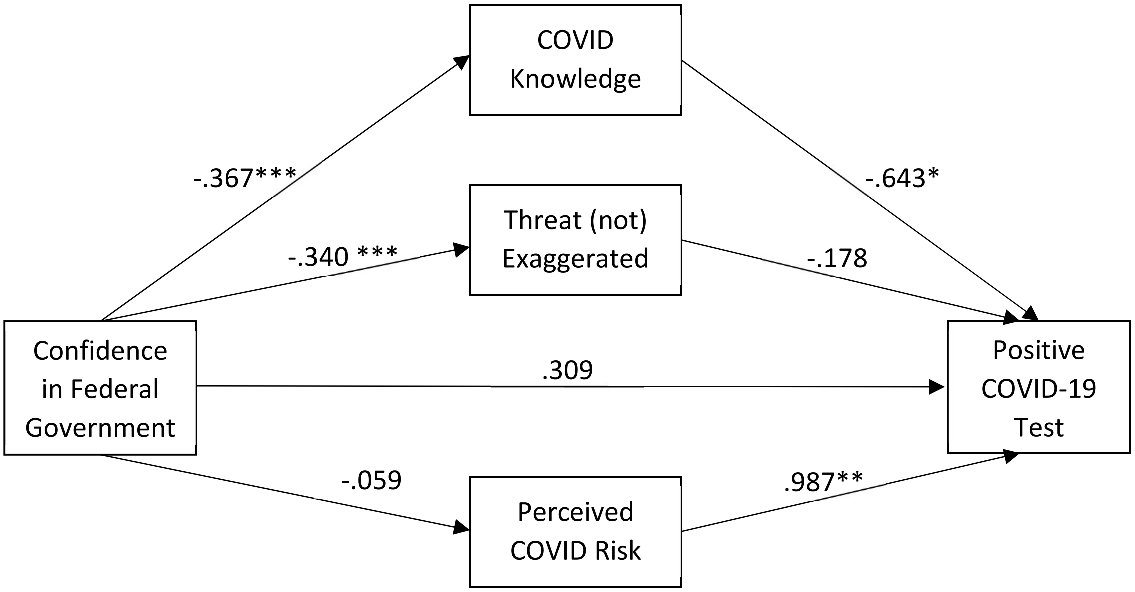


*$p < .05$; **$p < .01$; ***$p < .001$

*Supplementary Table 3. Indirect effects of confidence in federal government (re COVID-19 crisis) on COVID-19 test results*

| | Indirect Effect | SE | 95% Confidence Interval |
|---|---|---|---|
| TOTAL | .238 | .131 | [-.016, .508] |
| COVID Knowledge | .236† | .111 | [.049, .491] |
| Threat (not) Exaggerated | .061 | .133 | [-.220, .309] |
| Perceived COVID Risk | -.058 | .057 | [-.195, .028] |

†denotes a 95% confidence interval that does not include zero

*Supplementary Figure 4. Direct effects of general confidence in Trump on COVID-19 test results mediated by COVID-specific predictors*

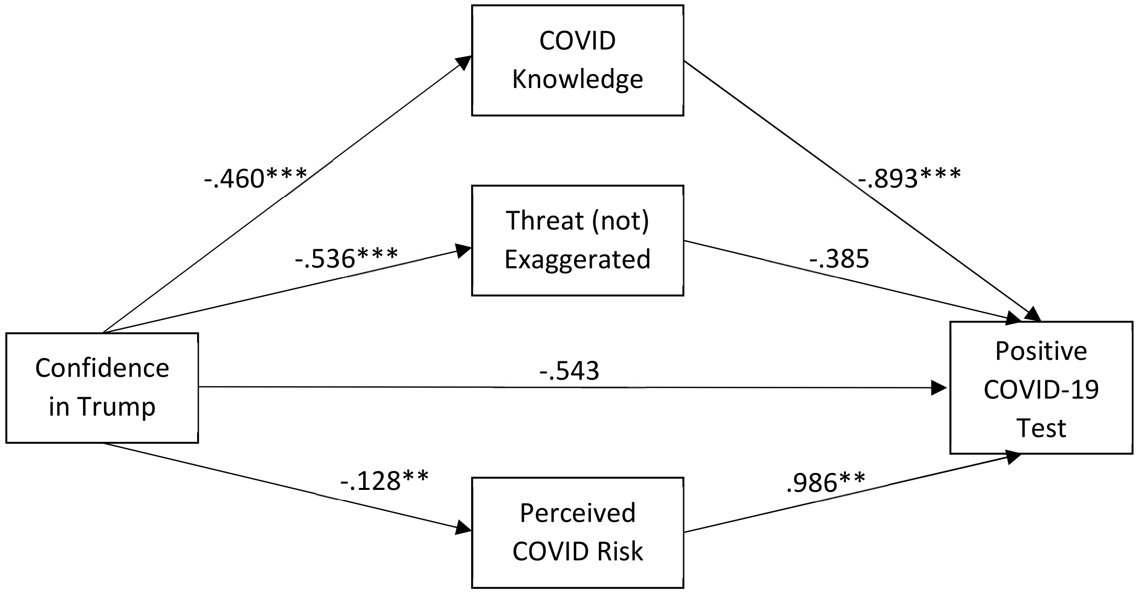


*$p < .05$; **$p < .01$; ***$p < .001$

*Supplementary Table 4. Indirect effects of general confidence in Trump on COVID-19 test results*

| | Indirect Effect | SE | 95% Confidence Interval |
|---|---|---|---|
| TOTAL | .491† | .197 | [.117, .910] |
| COVID Knowledge | .410† | .147 | [.166, .747] |
| Threat (not) Exaggerated | .206 | .214 | [-.208, .638] |
| Perceived COVID Risk | -.126† | .075 | [-.316, -.026] |

†denotes a 95% confidence interval that does not include zero

*Supplementary Figure 5. Direct effects of general interpersonal compassion on COVID-19 test results mediated by COVID-specific predictors*

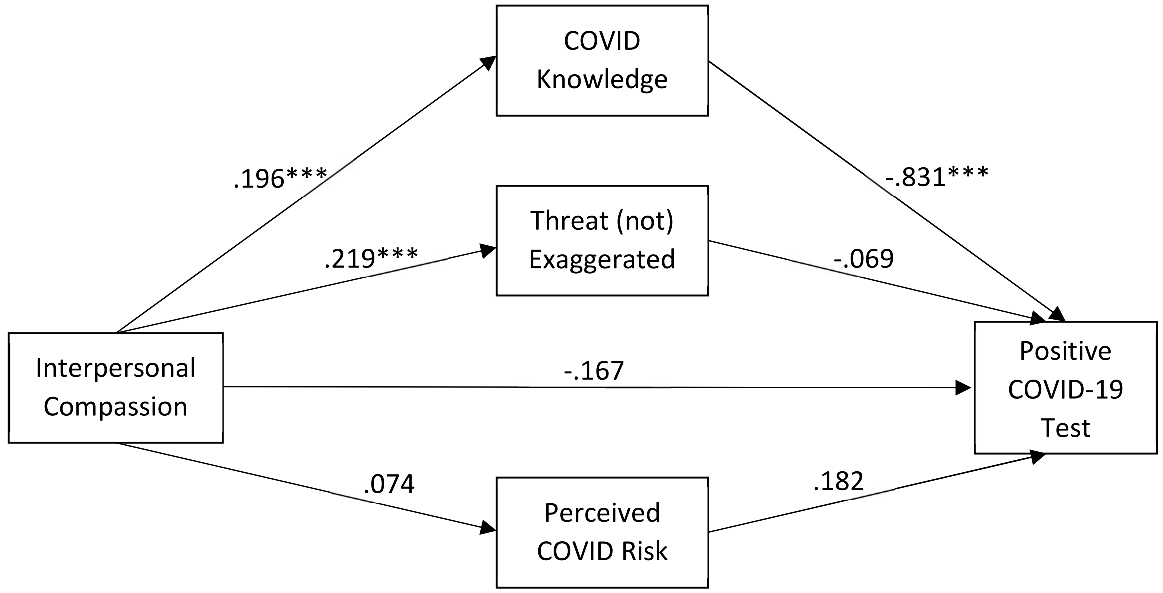


*$p < .05$; **$p < .01$; ***$p < .001$

*Supplementary Table 5. Indirect effects of general interpersonal compassion on COVID-19 test results*

| | Indirect Effect | SE | 95% Confidence Interval |
|---|---|---|---|
| TOTAL | -.165 † | .079 | [-.335, -.021] |
| COVID Knowledge | -.163† | .049 | [-.278, -.087] |
| Threat (not) Exaggerated | -.015 | .073 | [-.160, .133] |
| Perceived COVID Risk | .013 | .024 | [-.026, .073] |

†denotes a 95% confidence interval that does not include zero

*Supplementary Figure 6. Direct effects of disgust sensitivity on COVID-19 test results mediated by COVID-specific predictors*

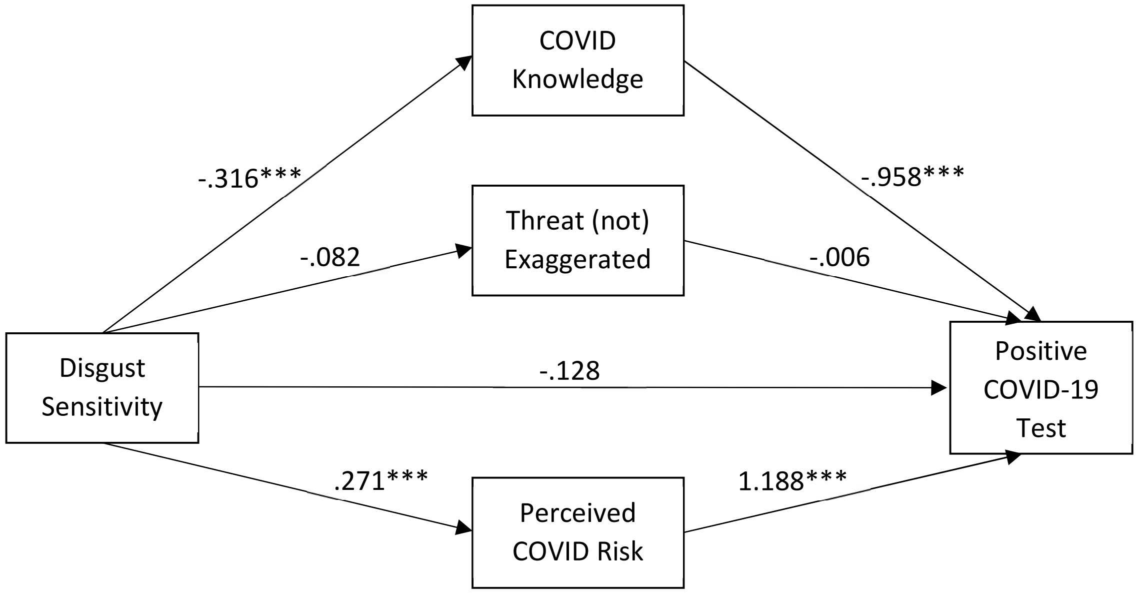


*$p < .05$; **$p < .01$; ***$p < .001$

*Supplementary Table 6. Indirect effects of disgust sensitivity on COVID-19 test results*

| | Indirect Effect | SE | 95% Confidence Interval |
|---|---|---|---|
| TOTAL | .628† | .146 | [.425, 1.010] |
| COVID Knowledge | .306† | .076 | [.193, .494] |
| Threat (not) Exaggerated | .001 | .026 | [-.055, .057] |
| Perceived COVID Risk | .321† | .122 | [.145, .622] |

†denotes a 95% confidence interval that does not include zero

*Supplementary Figure 7. Direct effects of perceived vulnerability to disease on COVID-19 test results mediated by COVID-specific predictors*

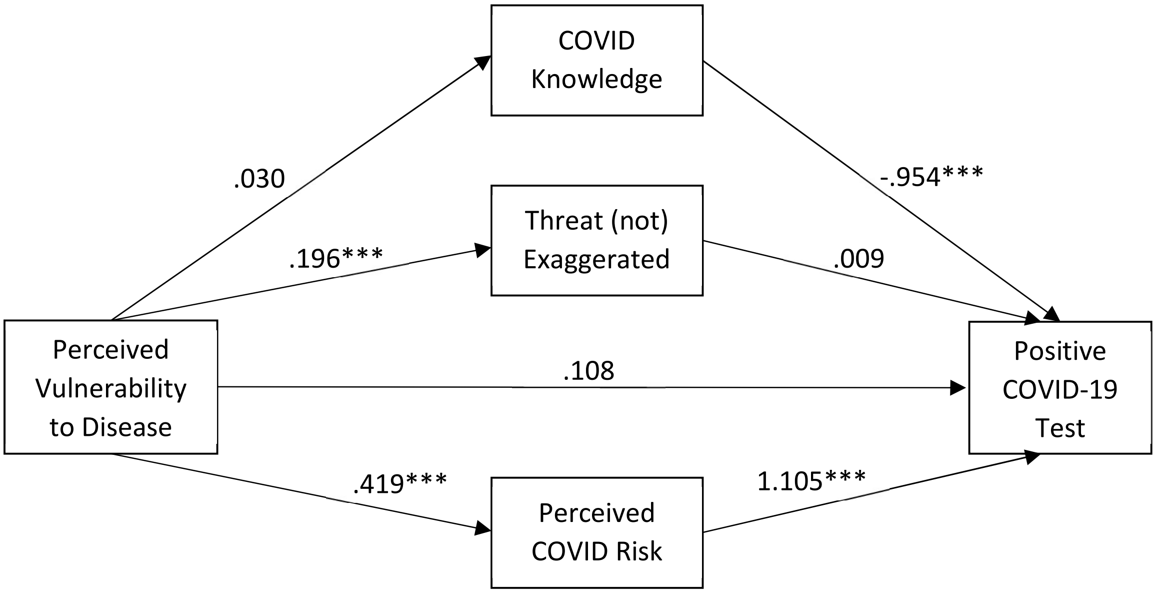


*$p < .05$; **$p < .01$; ***$p < .001$

*Supplementary Table 7. Indirect effects of perceived vulnerability to disease on COVID-19 test results*

| | Indirect Effect | SE | 95% Confidence Interval |
|---|---|---|---|
| TOTAL | .437† | .180 | [.163, .871] |
| COVID Knowledge | -.028 | .028 | [-.087, .023] |
| Threat (not) Exaggerated | .002 | .055 | [-.108, .116] |
| Perceived COVID Risk | .464† | .170 | [.213, .874] |

†denotes a 95% confidence interval that does not include zero

*Supplementary Figure 8. Direct effects of preexisting conditions on COVID-19 test results mediated by COVID-specific predictors*

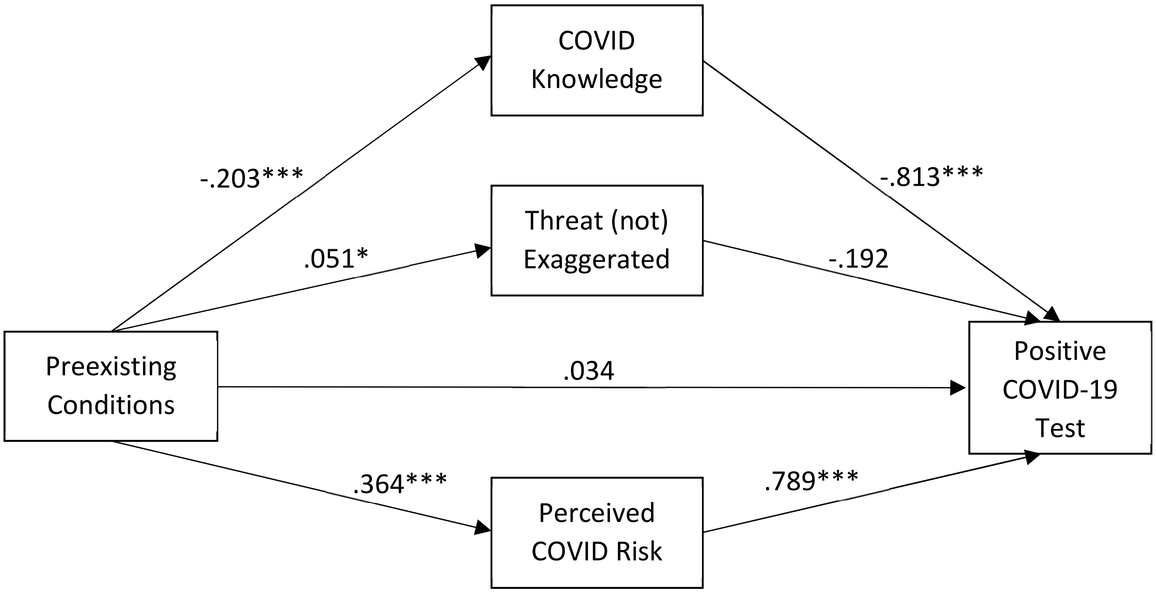


**p* < .05; ***p* < .01; ****p* < .001

*Supplementary Table 8. Indirect effects of preexisting conditions on COVID-19 test results*

| | Indirect Effect | SE | 95% Confidence Interval |
|---|---|---|---|
| TOTAL | .443† | .074 | [.317, .605] |
| COVID Knowledge | .165† | .028 | [.115, .227] |
| Threat (not) Exaggerated | -.010 | .010 | [-.032, .006] |
| Perceived COVID Risk | .287† | .070 | [.163, .438] |

†denotes a 95% confidence interval that does not include zero

*Supplementary Figure 9. Direct effects of political ideology on COVID-19 test results mediated by COVID-specific predictors*

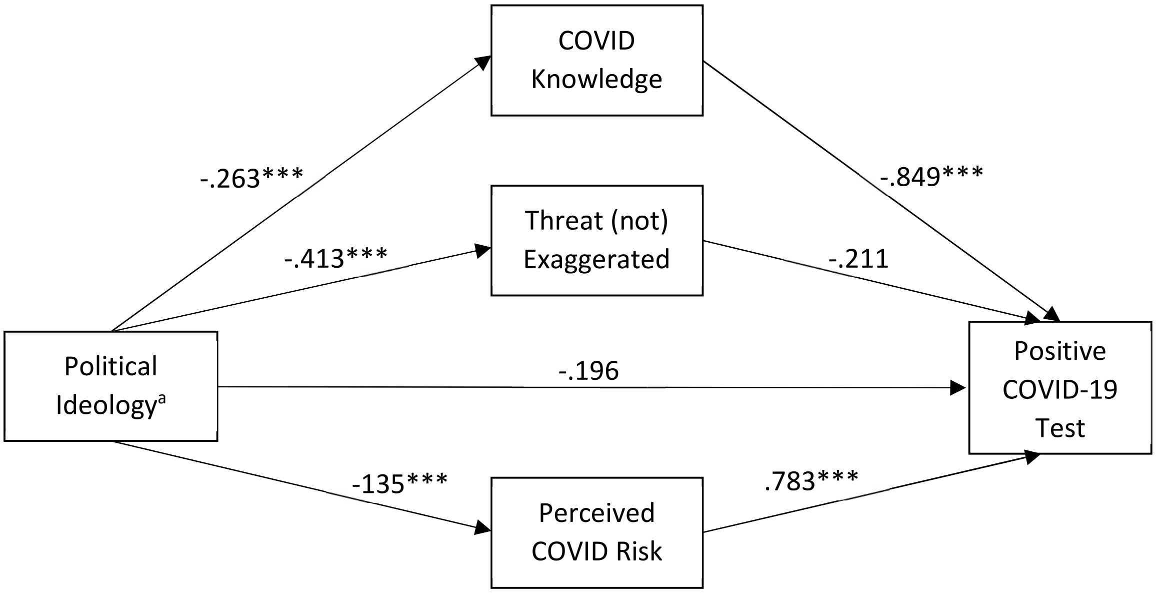


[a] Scale from 1 (Extremely liberal) to 7 (Extremely conservative)
**p* < .05; ***p* < .01; ****p* < .001

*Supplementary Table 9. Indirect effects of political ideology on COVID-19 test results*

| | Indirect Effect | SE | 95% Confidence Interval |
|---|---|---|---|
| TOTAL | .205† | .067 | [.072, .336] |
| COVID Knowledge | .224† | .034 | [.162, .298] |
| Threat (not) Exaggerated | .087 | .068 | [-.048, .221] |
| Perceived COVID Risk | -.106 † | .032 | [-.178, -.054] |

†denotes a 95% confidence interval that does not include zero

*Supplementary Figure 10. Direct effects of conspiratorial ideation on COVID-19 test results mediated by COVID-specific predictors*

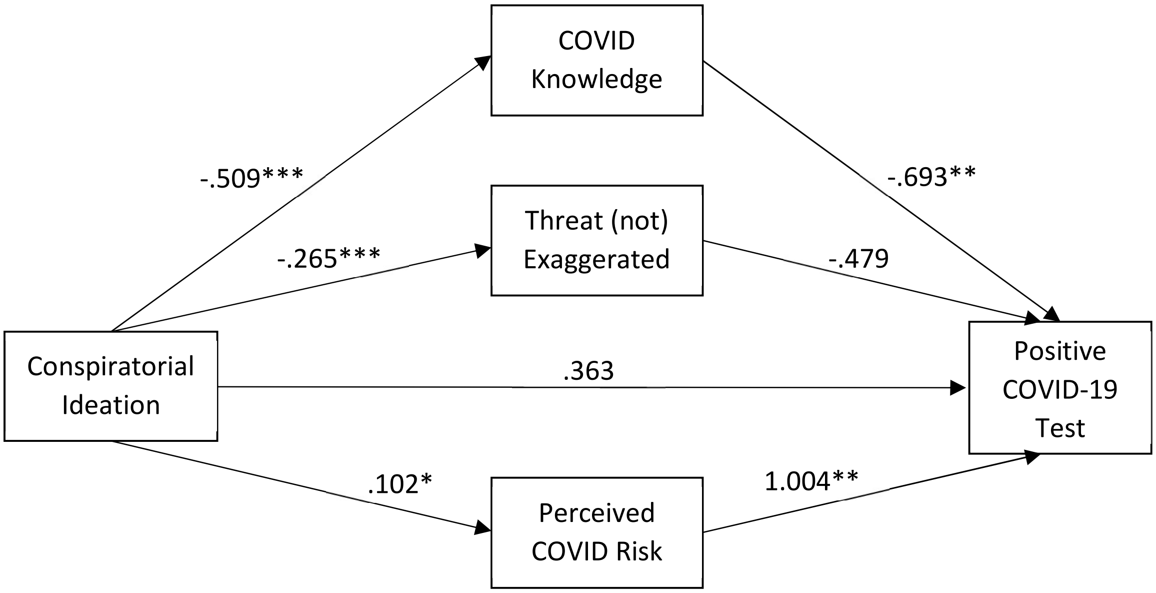


**p* < .05; ***p* < .01; ****p* < .001

*Supplementary Table 10. Indirect effects of conspiratorial ideation on COVID-19 test results*

| | Indirect Effect | SE | 95% Confidence Interval |
|---|---|---|---|
| TOTAL | .582† | .143 | [.366, .932] |
| COVID Knowledge | .353† | .133 | [.132, .665] |
| Threat (not) Exaggerated | .127 | .099 | [-.057, .339] |
| Perceived COVID Risk | .102† | .063 | [.009, .254] |

†denotes a 95% confidence interval that does not include zero

*Supplementary Figure 11. Direct effects of science literacy on COVID-19 test results mediated by COVID-specific predictors*

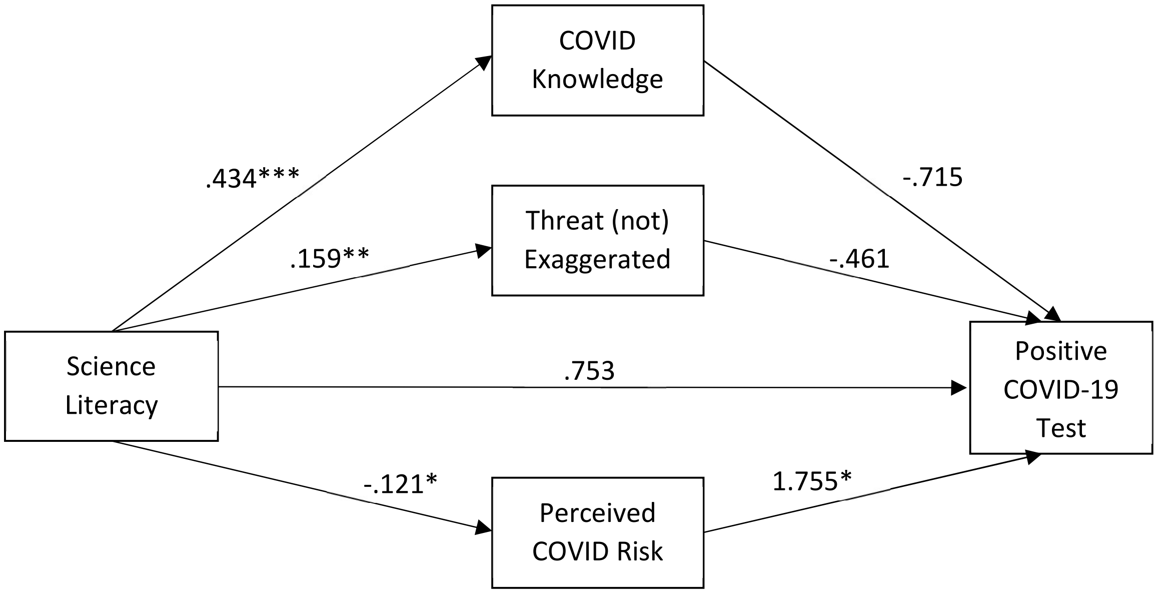


*$p$ < .05; **$p$ < .01; ***$p$ < .001

Note: The science literacy scale was included only in Study 1 at Time 1. Of the 304 participants who were responded to this measure, only 5 had positive COVID-19 tests. The bootstrapped mediational model failed to converge. Hence, the estimates should be interpreted cautiously.

*Supplementary Table 11. Indirect effects of science literacy on COVID-19 test results*

| | Indirect Effect | SE | 95% Confidence Interval |
|---|---|---|---|
| TOTAL | -.595† | 295,828.911 | [-413.573, -.186] |
| COVID Knowledge | -.310 | 111,285.505 | [-136.126, .322] |
| Threat (not) Exaggerated | -.073 | 88,852.714 | [-104.762, .361] |
| Perceived COVID Risk | -.212 | 122,323.743 | [-166.759, .019] |

†denotes a 95% confidence interval that does not include zero

Note: The science literacy scale was included only in Study 1 at Time 1. Of the 304 participants who were administered this measure, only 5 had positive COVID-19 tests. The bootstrapped mediational model failed to converge. Hence, the estimates should be interpreted cautiously.

*Supplementary Figure 12. Direct effects of Fox News use on COVID-19 test results mediated by COVID-specific predictors*

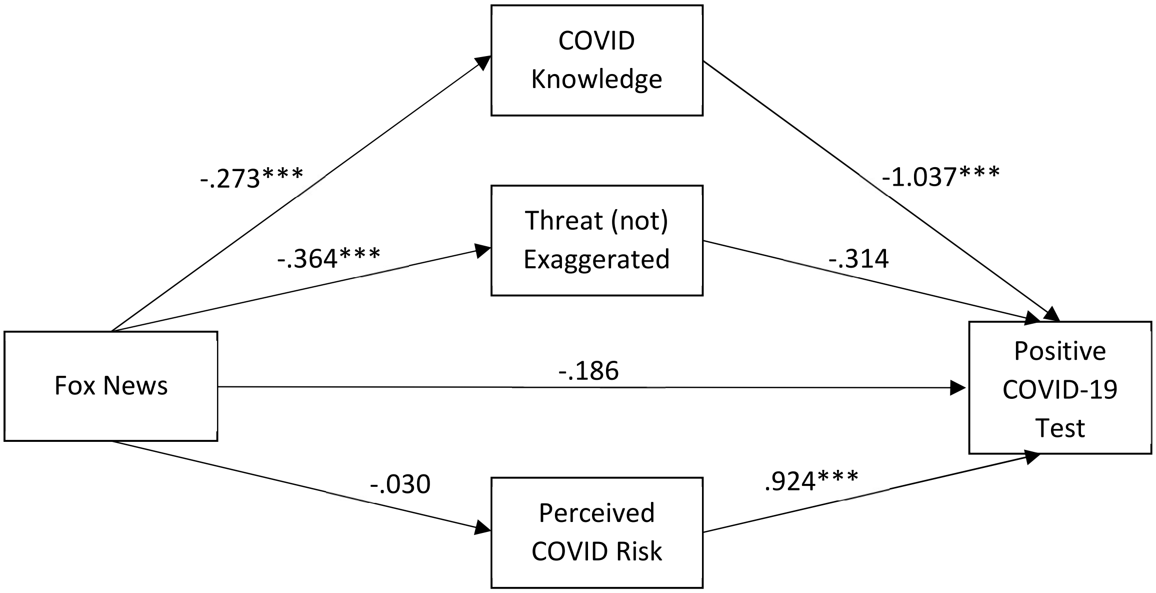


*$p < .05$; **$p < .01$; ***$p < .001$

*Supplementary Table 12. Indirect effects of Fox News use on COVID-19 test results*

| | Indirect Effect | SE | 95% Confidence Interval |
|---|---|---|---|
| TOTAL | .370† | .099 | [.189, .580] |
| COVID Knowledge | .284† | .065 | [.178, .433] |
| Threat (not) Exaggerated | .114 | .089 | [-.059, .298] |
| Perceived COVID Risk | -.028 | .046 | [-.123, .057] |

†denotes a 95% confidence interval that does not include zero

*Supplementary Figure 13. Direct effects of NPR use on COVID-19 test results mediated by COVID-specific predictors*

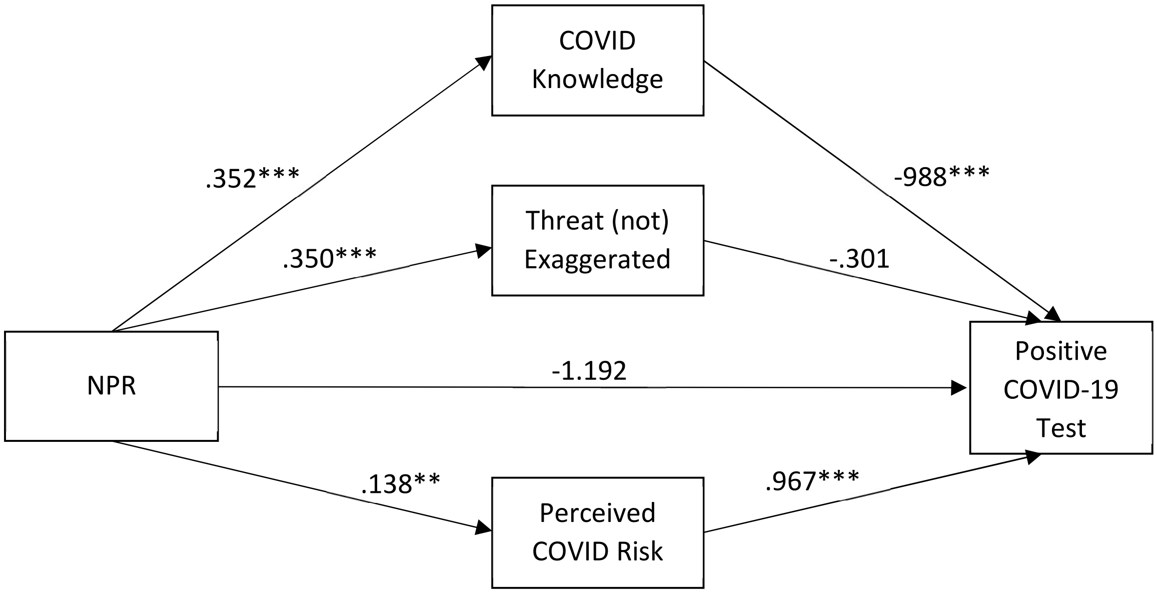


**p* < .05; ***p* < .01; ****p* < .001

*Supplementary Table 13. Indirect effects of NPR use on COVID-19 test results*

| | Indirect Effect | SE | 95% Confidence Interval |
|---|---|---|---|
| TOTAL | -.320† | .102 | [-.528, -.126] |
| COVID Knowledge | -.348† | .067 | [-.500, -.238] |
| Threat (not) Exaggerated | -.106 | .084 | [-.283, .050] |
| Perceived COVID Risk | -.134† | .064 | [.033, .285] |

†denotes a 95% confidence interval that does not include zero

*Supplementary Figure 14. Direct effects of national newspaper and/or magazine use on COVID-19 test results mediated by COVID-specific predictors*

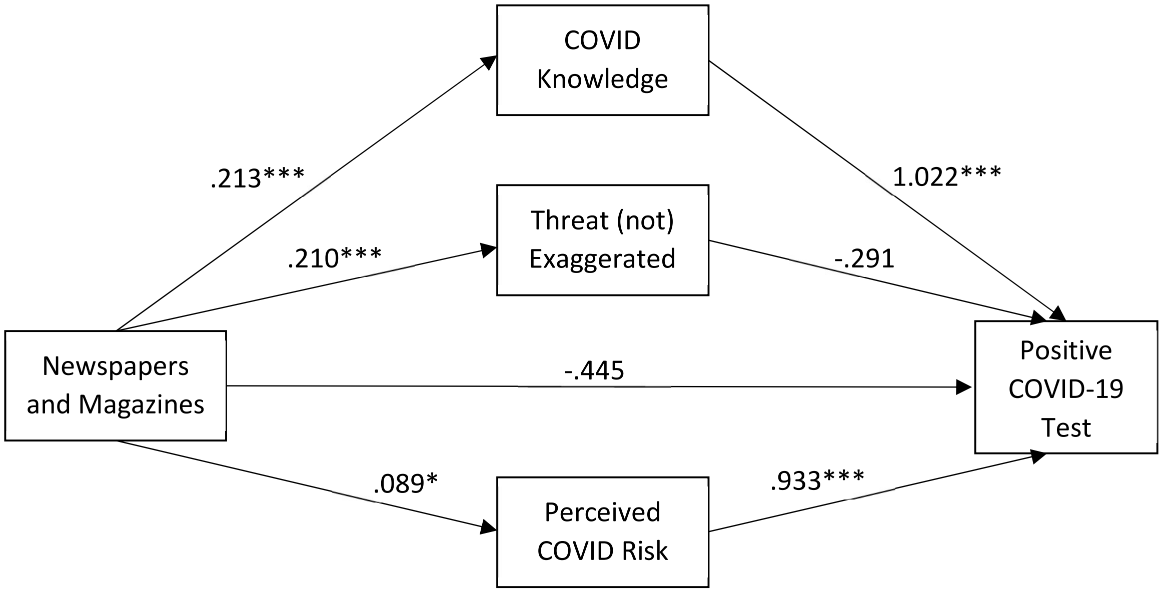


*$p < .05$; **$p < .01$; ***$p < .001$

*Supplementary Table 14. Indirect effects of national newspaper and/or magazine use on COVID-19 test results*

| | Indirect Effect | SE | 95% Confidence Interval |
|---|---|---|---|
| TOTAL | -.196† | .077 | [-.356, -.050] |
| COVID Knowledge | -.218† | .052 | [-.338, -.132] |
| Threat (not) Exaggerated | -.061 | .051 | [-.168, .035] |
| Perceived COVID Risk | .083† | .044 | [.010, .185] |

†denotes a 95% confidence interval that does not include zero

*Supplementary Figure 15. Direct effects of age on COVID-19 test results mediated by COVID-specific predictors*

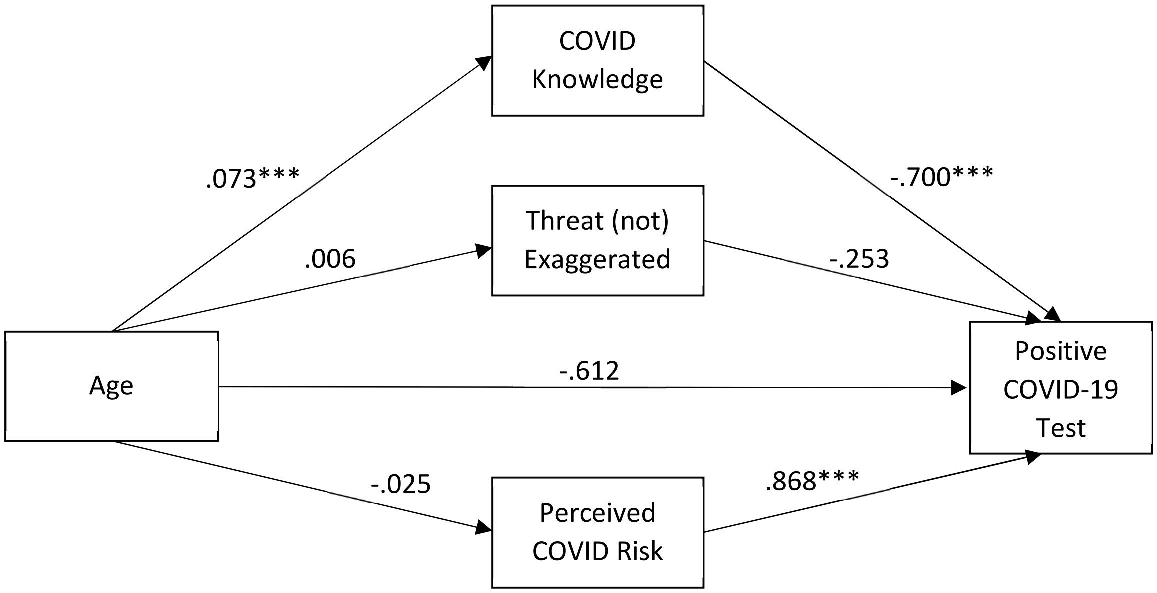


*$p < .05$; **$p < .01$; ***$p < .001$

*Supplementary Table 15. Indirect effects of age on COVID-19 test results*

| | Indirect Effect | SE | 95% Confidence Interval |
|---|---|---|---|
| TOTAL | -.074† | .043 | [-.176, -.019] |
| COVID Knowledge | -.051† | .028 | [-.117, -.016] |
| Threat (not) Exaggerated | -.002 | .007 | [-.022, .009] |
| Perceived COVID Risk | -.022 | .020 | [-.073, .004] |

†denotes a 95% confidence interval that does not include zero

*Supplementary Figure 16. Direct effects of gender on COVID-19 test results mediated by COVID-specific predictors*

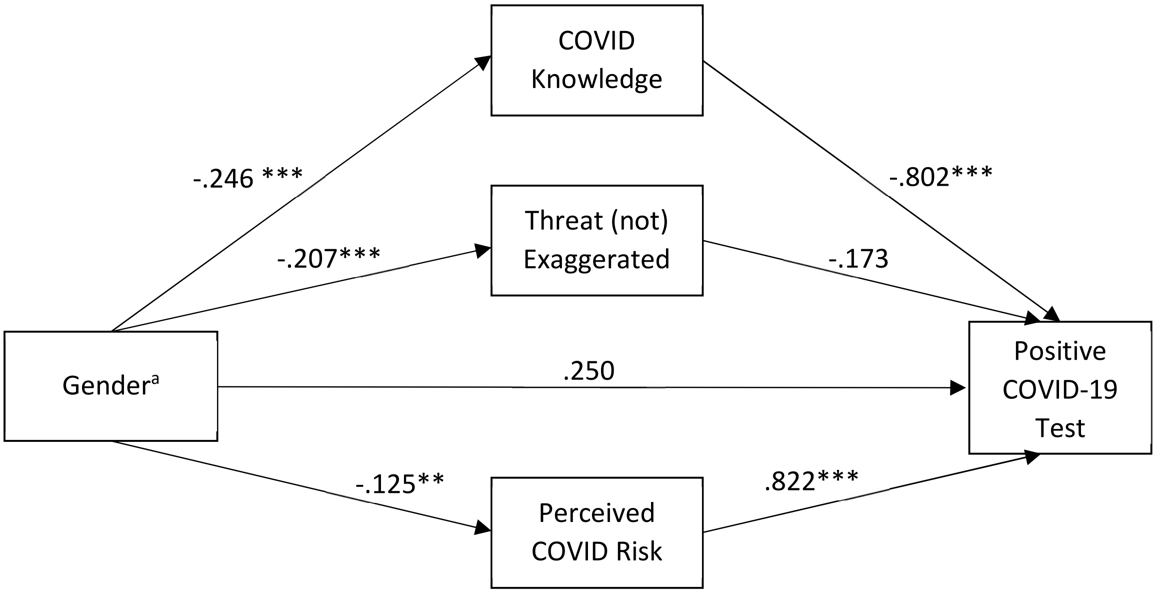


[a] Coded 1=male and 0=female; “Other” and “Prefer not to answer” responses were coded as missing for ease of interpretation

*$p < .05$; **$p < .01$; ***$p < .001$

*Supplementary Table 16. Indirect effects of gender on COVID-19 test results*

| | Indirect Effect | SE | 95% Confidence Interval |
|---|---|---|---|
| TOTAL | -.131 | .068 | [-.005, .262] |
| COVID Knowledge | .198[†] | .043 | [.122, .289] |
| Threat (not) Exaggerated | .036 | .036 | [-.034, .110] |
| Perceived COVID Risk | -.102[†] | .044 | [-.204, -.029] |

[†] denotes a 95% confidence interval that does not include zero

*Supplementary Figure 17. Direct effects of race on COVID-19 test results mediated by COVID-specific predictors*

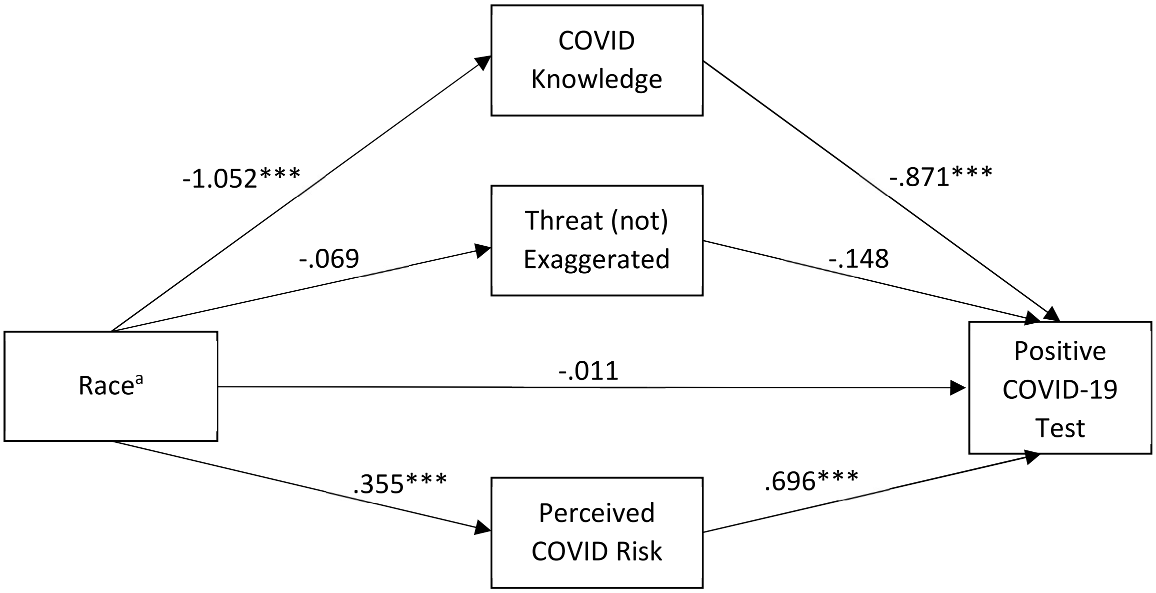


[a] Coded 1=Black and 0=White; all other racial and ethnic categories were coded as missing for ease of interpretation
**p* < .05; ***p* < .01; ****p* < .001

*Supplementary Table 17. Indirect effects of race on COVID-19 test results*

| | Indirect Effect | SE | 95% Confidence Interval |
|---|---|---|---|
| TOTAL | 1.173† | .184 | [.849, 1.563] |
| COVID Knowledge | .916† | .154 | [.648, 1.257] |
| Threat (not) Exaggerated | .010 | .021 | [-.025, .062] |
| Perceived COVID Risk | .247† | .090 | [.103, .455] |

†denotes a 95% confidence interval that does not include zero
